# MedMLX: On-Device Medical Imaging AI for Apple Silicon

**DOI:** 10.64898/2026.10.28.26366762

**Authors:** Joseph Sandoval, Alfredo Sandoval, Arsalan Saleem

## Abstract

**Background:** Medical imaging research increasingly combines specialized artificial intelligence models for segmentation, detection, image synthesis and report generation. For three-dimensional CT and MRI, these workflows typically run on NVIDIA GPUs with substantial memory. On-device AI on widely available Apple Silicon computers could broaden access to these models for clinical researchers. These computers share a large memory pool between the CPU and GPU, which suits workflows that combine several models. However, few established imaging models run natively on Apple Silicon. Adapting a model to new hardware also requires verifying that its results match the original software.

**Methods:** We developed MedMLX, a collection of 21 established medical imaging models rewritten for Apple Silicon in MLX, Apple’s machine learning framework. The collection includes nnU-Net, TotalSegmentator, MedSAM2 and SAM 3. The models segment organs and tumors, detect lung nodules, generate synthetic images and draft radiology reports from CT, MRI, PET/CT, radiographs and endoscopic images. Using public cases from 11 imaging datasets, we compared the models’ results with those of the original software run on an Intel CPU and an NVIDIA GPU. Agreement required a Dice coefficient of at least 0.99 for every segmentation label and equivalent criteria for other outputs. We measured memory use and processing time on an Apple M1 Max computer with 64 GB of memory.

**Results:** MedMLX agreed with the original software in 211 of 212 comparisons. The single exception was a brain MRI segmentation. On this case, the original software itself gave different results on the two devices. We illustrated sequential workflows, including anatomical segmentation of a synthetic brain MRI and lesion synthesis guided by organ segmentation. With a memory-saving setting, 19 of the 21 models used less than 16 GB of memory. This setting reduced memory use by a median of 49% and increased processing time by a median of 13%.

**Conclusions:** MedMLX enables on-device AI with established medical imaging models on Apple Silicon computers rather than specialized NVIDIA hardware. The models agree closely with the original software, and most need less than 16 GB of memory.

## Introduction

AI-assisted medical imaging workflows draw on specialized models for complementary tasks, including segmentation [1], detection [2], classification, image synthesis [3], [4] and report generation [5]. A single study may involve several models operating on the same images or on outputs produced earlier in the workflow. When several models remain in memory for sequential or concurrent use, memory capacity and resource management become important, particularly for volumetric images.

Medical imaging frameworks such as nnU-Net [1] and MONAI [6] are developed primarily for NVIDIA GPUs and CUDA. Mac computers offer a widely available alternative: macOS runs on about one in six desktop computers worldwide [7], and a third of software developers responding to a 2025 survey used it for their work [8]. On-device AI, in which models run on the user’s own computer, could broaden access to imaging AI research for clinical researchers without specialized NVIDIA hardware. Running models on device also means that imaging data need not be transferred to shared or cloud computing services.

Apple Silicon computers, which include all current Macs, combine GPU acceleration with a large unified memory pool shared by the CPU and GPU. MLX is a native machine learning framework in which both processors operate on the same arrays without transfers between separate memory pools [9]. These capabilities suit workflows that keep several models, image volumes and intermediate results in memory. Most medical imaging models, however, are released for PyTorch [10] or TensorFlow, and each depends on model-specific preprocessing, inference settings and postprocessing. A native SAM 2.1 implementation [11] and medical tools using PyTorch’s Metal backend [12] support local image analysis on these computers; extending this support across tasks requires preserving the behavior of each released model.

Here, we introduce MedMLX, which makes established medical imaging models available for on-device AI on Apple Silicon. The collection comprises 21 pretrained models from 16 architecture families implemented natively in MLX. The models support segmentation, detection, image synthesis, report generation and feature extraction, including text-prompted segmentation with SAM 3 [13]. Each package includes model-specific preprocessing and postprocessing, with a shared Python API, command-line interface and local Model Context Protocol server.

Moving released models to a new hardware and software stack requires checking that their behavior is preserved, because differences in numerical precision, accumulation order and preprocessing can change the final predictions. We therefore first assessed output agreement between MedMLX and the original implementations on Apple Silicon, Intel CPU and NVIDIA GPU hardware. We then measured the memory and runtime needed to run each model on device, including the effect of the MLX cache, and compared network runtime in MLX, Core ML and Core AI on M1 Max and M5 Max computers.

### Software design

Each package combines a pretrained network with the preprocessing and postprocessing used by its original implementation. Network inference runs in MLX on the Apple GPU; CPU libraries handle image reading, preprocessing and export. Segmentation masks are mapped back to the input image grid. Packages share network code where appropriate: the nnU-Net implementation supports multiple organ and lesion models [1], [14], [15], and the brain tumor and renal models share SegResNet code [16].

Every package provides a Python `run()` function. The `medmlx` package adds a common Python API, command-line interface and Model Context Protocol (MCP) server for listing models and checkpoints, inspecting arguments, checking inputs and weights, and running models. The local MCP server allows compatible clients, including AI coding assistants, to run the same model operations. Its execution and job handling are described in Supplementary Note 1.

Model-specific operators reproduce the original rounding and accumulation behavior where these affect output agreement. Optional profiles for renal SegResNet and SynthSeg 2.0 use higher-precision arithmetic to reduce numerical differences across devices (Methods).

The 21 evaluated models cover segmentation, detection, image synthesis, report generation and feature extraction across CT, MRI, PET/CT, radiography and endoscopy (Table 1). Figure 1 shows example outputs and a sequential workflow in which SynthSeg 2.0 segments and parcellates a T1 MRI generated by NV-Generate-MR-Brain (panels n–p). A second workflow uses TotalSegmentator masks as inputs to LesionDiffusion (Supplementary Fig. S12). Supplementary Figure S7 shows an NV-Reason-CT report alongside its source CT.

**Fig. 1.**
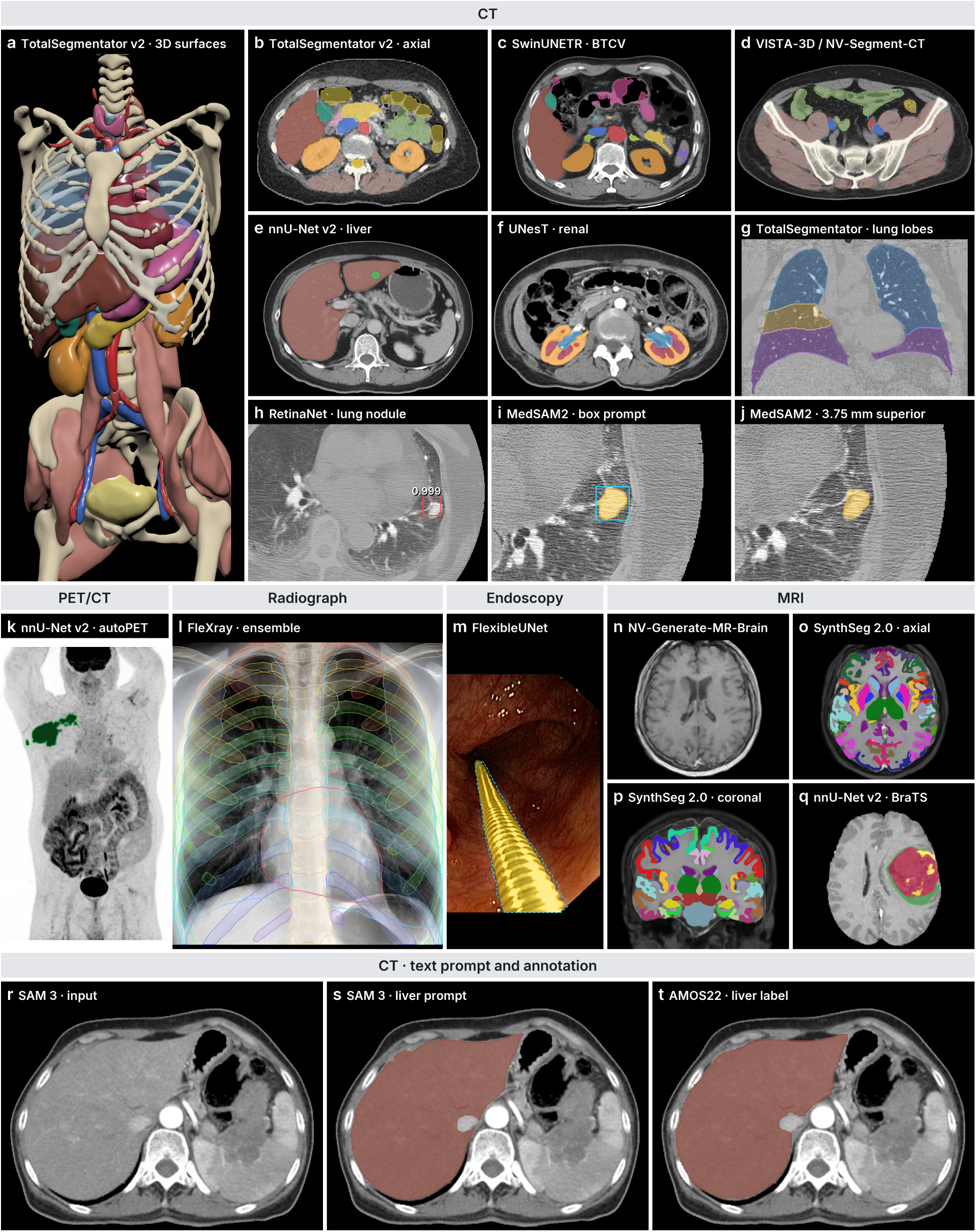
Example MedMLX outputs. (a, b) TotalSegmentator [17] fast pipeline on an autoPET CT, shown as surfaces and an axial slice. (c) SwinUNETR abdominal organ segmentation. (d) VISTA-3D segmentation of all 117 classes. (e) nnU-Net liver and liver tumor segmentation. (f) UNesT renal substructures. (g) TotalSegmentator lung lobes. (h) RetinaNet lung nodule detections. (i, j) MedSAM2 lung nodule segmentation from a box prompt, on the prompted slice and 3.75 mm superior to it. (k) nnU-Net FDG-avid lesion segmentation on a PET maximum-intensity projection. (l) Chest radiograph structures segmented by the five-model FleXray ensemble. (m) FlexibleUNet endoscopic instrument segmentation; the dashed outline marks the reference mask. (n) NV-Generate synthetic T1 MRI. (o, p) SynthSeg 2.0 segmentation and parcellation of the image in n. (q) nnU-Net glioma segmentation. (r–t) AMOS22 CT slice with the largest annotated liver area, SAM 3 output for the text prompt “liver” at confidence 0.5, and the AMOS22 liver annotation. This case was selected because SAM 3 produced a liver proposal. Panels e, k and q were selected for the highest whole-volume Dice against annotations among five, three and five candidates, respectively; panel f was selected for renal contrast enhancement. Supplementary Note 3 describes comparisons of the complete TotalSegmentator pipeline, including the lung lobes in g; Supplementary Note 5 reports the FleXray ensemble comparisons.

## Results

### Agreement with the original implementations

Across public imaging cases from 11 datasets, MLX outputs were identical to, or within tolerance of, the original outputs in 211 of 212 comparisons (Fig. 2; Supplementary Table S7). These comparisons covered 20 of the 21 models, plus the FleXray ensemble and conditioned LesionDiffusion, with matched random noise for generative models. The single comparison outside tolerance was SynthSeg 2.0 on BRATS_004: the MLX output differed from the Intel CPU output beyond tolerance but agreed with the RTX 4090 output. The original Intel CPU and RTX 4090 outputs also differed beyond tolerance on this case.

BioMedParse v2 [18], [19] was compared on a 63-slice AMOS22 CT. With matched attention sampling, FP32 MLX on the M1 Max GPU and the original PyTorch implementation on the same computer’s CPU produced identical spleen and right-kidney labels (Supplementary Table S3).

With higher-precision arithmetic in MLX and in the original implementations, renal SegResNet and SynthSeg 2.0 produced identical masks across all three devices for the five cases tested per model (Supplementary Note 5). On the M5 Max, standard FP32 renal SegResNet also produced identical masks in MLX on the GPU and in MONAI on the CPU for all five renal cases.

With matched random noise, LesionDiffusion produced identical lesion masks in MLX and the original implementation. Mean absolute differences between the generated CT images were below 1 HU both inside and outside the lesion masks, although local differences outside the masks reached 428 HU (Supplementary Table S6).

**Fig. 2.**
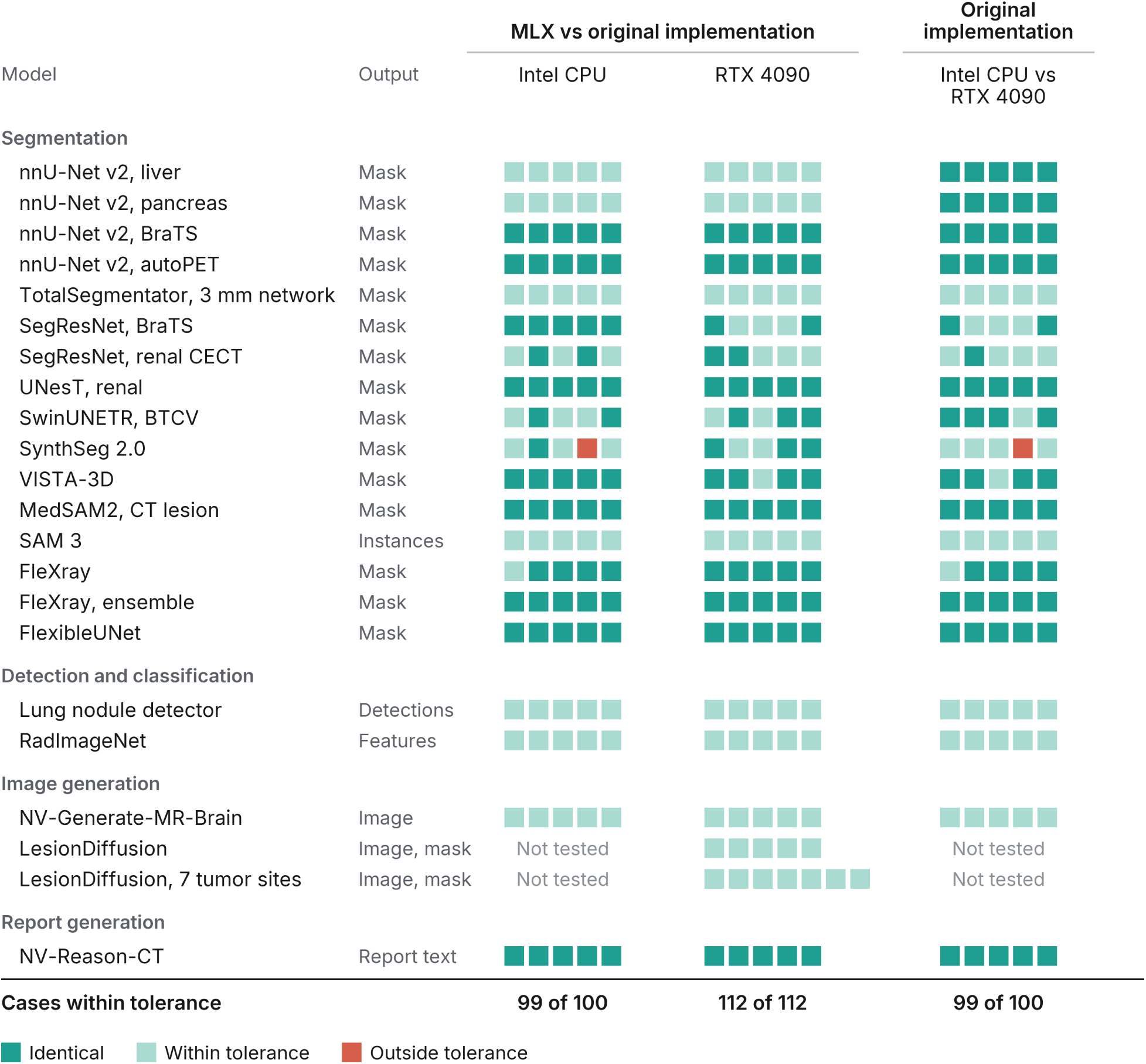
Output agreement between MLX and the original implementations with standard arithmetic. Each square represents one comparison on one case or generation seed. Rows contain five cases or seeds, except conditioned LesionDiffusion, which uses one organ crop for each of seven tumor sites. The left two columns compare MLX on the M1 Max GPU with the original implementation on the Intel CPU and RTX 4090. The right column compares the two reference devices. Dark teal indicates identical masks or generated text; light teal indicates agreement within tolerance; red indicates a comparison outside tolerance. Tolerances are a Dice score of at least 0.99 for every mask label, cosine similarity of at least 0.9999 for images and features, and the detection and instance criteria given in Methods. LesionDiffusion was compared with the RTX 4090 only. The TotalSegmentator row evaluates the 3 mm network; complete-pipeline comparisons are in Supplementary Note 3. Some images were reused across seeds or organ crops. BioMedParse v2 was compared on one case (Supplementary Table S3).

### Memory requirements and package runtime

On the 64 GB M1 Max, disabling the MLX cache reduced peak memory by a median of 49% across models, with a median 13% increase in package runtime (Fig. 3; Supplementary Table S8). With the cache disabled, median peak memory ranged from 0.3 to 27.6 GB and was below 16 GB for 19 of the 21 models, compared with 13 with the default cache. Median package runtimes ranged from 0.2 s (RadImageNet) to 1,353 s (LesionDiffusion) (Table 1).

Disabling the cache lowered peak memory for nnU-Net liver from 17.8 to 6.1 GB, SwinUNETR from 28.9 to 11.9 GB, NV-Generate from 53.0 to 27.6 GB and NV-Reason-CT from 53.5 to 23.3 GB. Its effect on runtime varied. NV-Generate took a median of 251 s with the default cache and 244 s with the cache disabled, whereas UNesT and SwinUNETR took 38% and 36% longer, respectively, with the cache disabled.

**Fig. 3.**
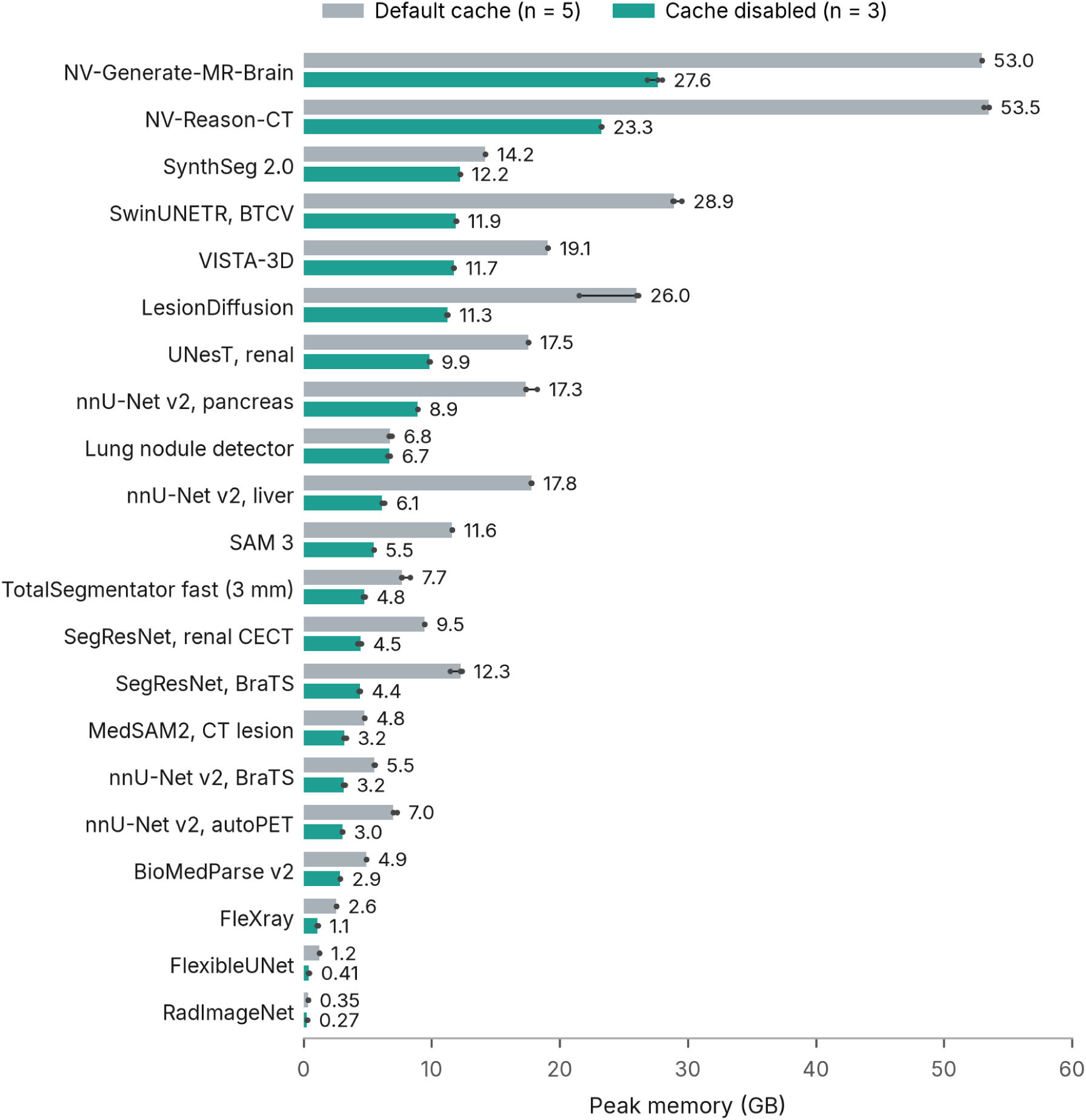
Peak memory with the default MLX cache and with the cache disabled on the 64 GB M1 Max. Bars and printed values show medians; dots show individual runs, and lines span their range. Each model ran five times with the default cache and three times with the cache disabled. Rows are ordered by median peak memory with the cache disabled. Workloads are defined in Table 1, and runtimes are reported in Supplementary Table S8.

**Table 1.** Package runtime and peak memory on the 64 GB M1 Max with the MLX cache disabled. Values are medians of three runs per model. Each row uses one input case (Supplementary Table S1). Results with the default cache are in Supplementary Table S8.

| Model | Workload | Time (s) | Peak memory (GB) |
| --- | --- | --- | --- |
| nnU-Net v2, liver | Liver and liver tumor segmentation in abdominal CT | 263.2 | 6.1 |
| nnU-Net v2, pancreas | Pancreas and pancreatic tumor segmentation in abdominal CT | 42.6 | 8.9 |
| nnU-Net v2, BraTS | Brain tumor segmentation in MRI | 4.9 | 3.2 |
| nnU-Net v2, autoPET | FDG-avid lesion segmentation in PET/CT | 16.0 | 3.0 |
| TotalSegmentator fast (3 mm) [17] | Segmentation of 117 structures in abdominal CT | 3.4 | 4.8 |
| SegResNet, BraTS | Brain tumor segmentation in MRI | 1.6 | 4.4 |
| SegResNet, renal CECT | Kidney, tumor, vessel and ureter segmentation in three-phase CT crops | 10.8 | 4.5 |
| UNesT, renal [20] | Renal cortex, medulla and pelvicalyceal system segmentation in abdominal CT | 144.1 | 9.9 |
| SwinUNETR, BTCV [21] | Segmentation of 13 abdominal organs in CT | 120.0 | 11.9 |
| SynthSeg 2.0 [22] | Brain segmentation and cortical parcellation in T1 MRI | 17.4 | 12.2 |
| VISTA-3D / NV-Segment-CT [23] | Anatomy segmentation in abdominal CT | 49.8 | 11.7 |
| FlexRay [24] | Lung segmentation in a chest radiograph | 1.1 | 1.1 |
| FlexibleUNet [25] | Instrument segmentation in an endoscopy frame | 0.2 | 0.4 |
| MedSAM2, CT lesion [26], [27] | Lung nodule segmentation from a box prompt in chest CT | 3.2 | 3.2 |
| SAM 3 [13] | Liver segmentation from a text prompt in one CT slice | 1.7 | 5.5 |
| BioMedParse v2 [18], [19] | Spleen and right kidney segmentation from text prompts in abdominal CT | 14.3 | 2.9 |
| Lung nodule detector [28] | Lung nodule detection in chest CT | 11.8 | 6.7 |
| NV-Generate-MR-Brain [3], [4] | Synthesis of a T1 brain MRI | 244.2 | 27.6 |
| LesionDiffusion [29] | Liver tumor synthesis in an abdominal CT crop | 1353.4 | 11.3 |
| NV-Reason-CT [5] | Structured report from chest CT | 84.8 | 23.3 |
| RadImageNet [30] | Feature extraction from one CT slice | 0.2 | 0.3 |

### Network runtime across execution frameworks

Fixed-input FP32 benchmarks compared 17 networks across MLX, Core ML [31] and Core AI [32] on the M1 Max and M5 Max (Fig. 4; Supplementary Table S9). After warm-up, MLX had the lowest median runtime for all 12 volumetric networks, and Core ML for all 5 two-dimensional networks, on both computers. The two-dimensional networks were RadImageNet, MedSAM2, FlexibleUNet, FleXray and SAM 3. Core AI did not have the lowest median runtime for any tested network. Across the volumetric networks, Core ML took a median of 1.80 times as long as MLX on the M1 Max and 1.93 times as long on the M5 Max.

**Fig. 4.**
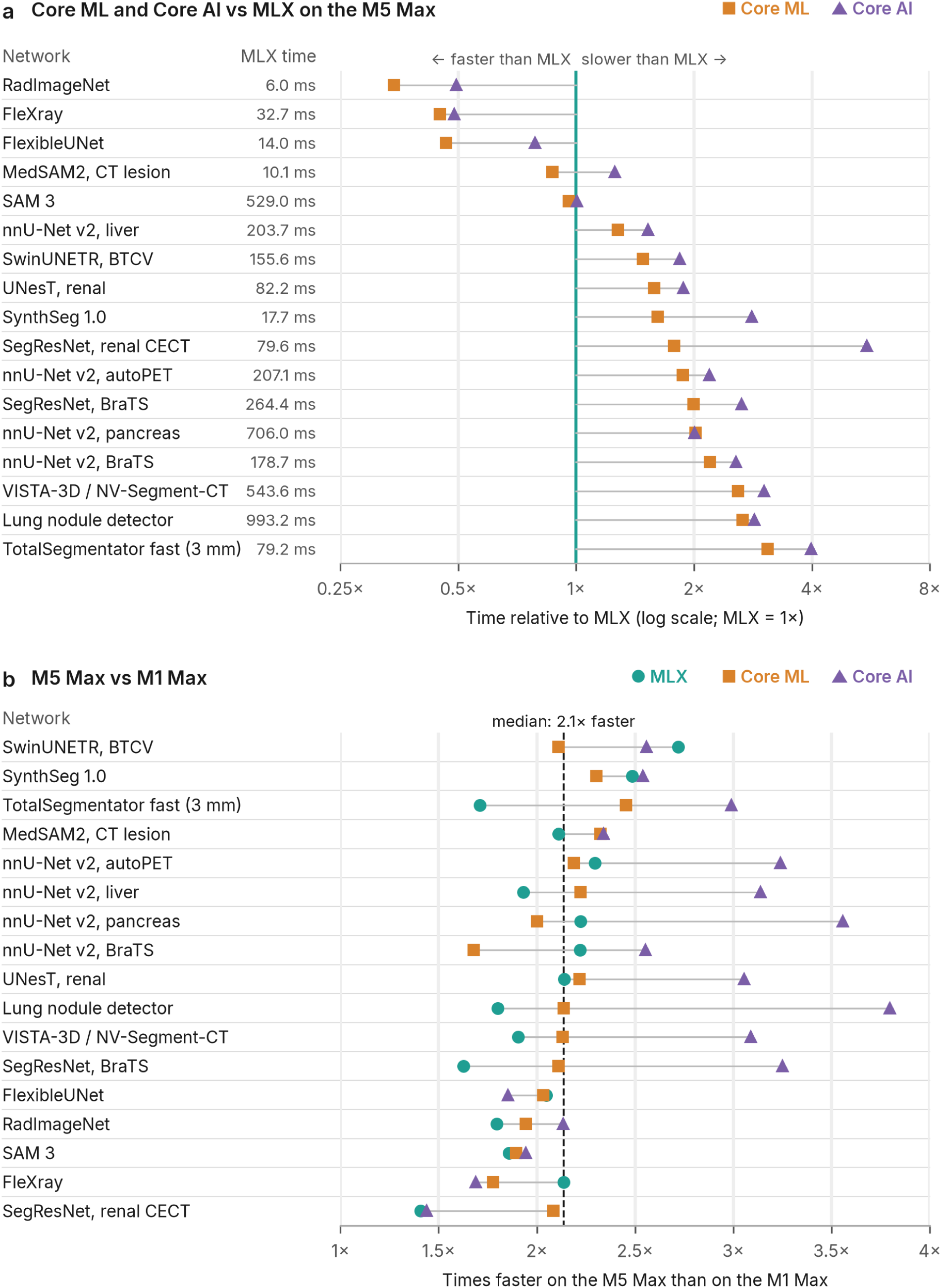
Network runtime across MLX, Core ML and Core AI. (a) Core ML and Core AI time divided by MLX time on the M5 Max, ordered by the Core ML ratio. Symbols to the left of the MLX reference line indicate shorter runtime than MLX. The MLX time column gives the MLX median on the M5 Max. (b) M1 Max time divided by M5 Max time for each network and runtime. Networks are ordered by their median speedup across the three runtimes; the dashed line marks the median across all network and runtime combinations. Each value is the median of five run medians, with ten timed calls per run after warm-up. All runtimes used FP32 and the same fixed input with batch size one; SAM 3 also received a 32-token prompt and attention mask. Timing began with the NumPy input and ended when the NumPy output was available. MLX used its GPU backend, Core ML used CPU_AND_GPU, and Core AI used its GPU profile. SynthSeg denotes the standalone 1.0 network. Supplementary Table S9 lists input sizes, times and ranges on both computers. Package runtime is reported separately in Table 1.

Across the tested network and runtime combinations, the M5 Max ran a median of 2.1 times faster than the M1 Max (range 1.4–3.8). Median speedups were 2.0 for MLX, 2.1 for Core ML and 2.6 for Core AI (Fig. 4b).

First-call performance differed from performance after warm-up. Excluding model loading, Core ML’s median first call was slower than MLX’s for all five two-dimensional networks on both computers. On the M5 Max, the first SAM 3 call took a median of 579 ms in MLX and 2,881 ms in Core ML, compared with 529 and 507 ms after warm-up. These network timings are separate from the package runtimes in Table 1.

## Discussion

MedMLX makes established medical imaging models available for on-device AI on Apple Silicon computers. Its outputs agreed closely with those of the original implementations on the selected inputs, and the memory and runtime measurements describe what is needed to run each model on device.

For clinical researchers with Apple Silicon computers, MedMLX provides a way to use established imaging models without specialized NVIDIA hardware or external computing services. Access depends on more than the networks themselves. Each package also includes the preprocessing and postprocessing of the original implementation, and the shared Python API, command-line interface and local MCP server give the packages a common entry point for scripts, command-line use and compatible AI assistants.

The agreement experiments assess whether the MLX implementations reproduce the behavior of the released models. Agreement can include shared errors: both UNesT implementations labeled the femoral heads as kidney on `amos_0008` (Supplementary Fig. S10), and all devices returned no liver proposal for two of the five SAM 3 slices at the fixed confidence threshold. These examples distinguish agreement between implementations from the anatomical accuracy of a prediction.

The only MLX comparison outside tolerance involved a SynthSeg 2.0 Intel CPU output that also disagreed with the original RTX 4090 output. Higher-precision arithmetic in MLX and in the original implementations removed the mask differences for SynthSeg 2.0 and renal SegResNet in the cases tested, consistent with floating-point rounding as the source of the remaining discrepancies. Because the arithmetic changed in both the MLX and original implementations, these profiles form a separate numerical comparison.

The MLX cache keeps freed memory for reuse [9], so the cache setting trades memory against runtime. On the 64 GB M1 Max, disabling the cache brought six of the eight models that exceeded 16 GB with the default cache below that level, while UNesT and SwinUNETR ran more than a third slower. Researchers can therefore choose the cache setting according to the memory of their computer and the processing time they can accept.

Runtime depended on both the network and how often it was called. MLX had the lowest median runtime after warm-up for the 12 volumetric networks tested, and Core ML for the 5 two-dimensional networks. For the two-dimensional networks, MLX was faster on the first call, whereas Core ML was faster after warm-up. For these models, the faster framework therefore depends on whether a model is called once or repeatedly, as in batch processing or interactive use. Package calls also include weight loading, preprocessing and export, so network timing alone does not determine the time needed to obtain a result.

Because each package includes its own preprocessing and postprocessing, the output of one model can serve as the input to another. Examples include anatomical segmentation of a synthetic brain MRI (Fig. 1n–p) and lesion synthesis guided by organ segmentation (Supplementary Fig. S12).

We measured models individually and illustrated sequential workflows; concurrent execution and sustained residency of several models were not benchmarked. The evaluation used selected public inputs, including some from dataset training partitions, and several comparisons reused images across generation seeds, organ crops or arithmetic profiles. Package measurements used one case per model on one M1 Max, and network benchmarks used one fixed synthetic input per network on one M1 Max and one M5 Max.

In conclusion, MedMLX enables on-device AI with established medical imaging models on Apple Silicon computers rather than specialized NVIDIA hardware. Its outputs agreed closely with those of the original implementations, and with the MLX cache disabled, most models required less than 16 GB of memory. The memory and runtime measurements can help researchers choose models and settings for their own computers.

## Methods

### Native implementations

We reimplemented each network in MLX from its original source code and loaded the released weights. Preprocessing, sliding-window settings and postprocessing followed the original implementation. We compared preprocessing, intermediate activations, network outputs and exported files with those of the original implementations, using seeded random weights with synthetic inputs and released checkpoints with full-size inputs.

### Evaluation design and input preparation

We evaluated each of the 21 models with its released weights on one example case or fixed generation seed (Supplementary Tables S1 and S3). Comparisons across multiple cases covered 20 models and the additional profiles listed in Supplementary Tables S5 and S7, using five cases or generation seeds per model or profile, except conditioned LesionDiffusion, which used one organ crop for each of seven tumor sites. We reported all selected cases, including those that failed the agreement criteria. Each run used a separate Python process.

Inputs were prepared in each model’s required format, spatial grid and intensity scale. Preparation included separating MRI channels, aligning CT and PET, selecting two-dimensional slices and extracting organ crops. Multiphase renal CT also required registration to the venous phase and denoising. Supplementary Note 2 describes the model-specific steps and parameters.

Inference followed the original implementation without optional test-time augmentation. nnU-Net models ran without mirroring and used one checkpoint per model, including the released fold_all pancreas checkpoint. For prompted and generative models, input boxes or text prompts, generation seeds and sampling or decoding settings are listed in Supplementary Table S1 and Supplementary Note 2.

We checked that every run finished and that its saved outputs could be read back. Images had the expected size and spacing, masks contained the expected labels, detections had valid boxes and scores, generated images had valid intensity values, reports had the expected sections and features had the expected length. We also viewed each 3D segmentation over its input image in axial, coronal and sagittal planes.

### Package runtime and memory measurements

Package runtime was the wall-clock time from the model call to completion of export. It included weight loading, the package’s preprocessing and inference, and excluded Python startup and model imports. Input preparation performed before the call, including renal registration, denoising and crop extraction, was outside the timed interval.

Each model ran three times with the MLX cache limit set to zero and five times with the default cache, each in a separate Python process without warm-up. The lung nodule detector sets its own 512 MiB cache limit in both settings. Table 1 reports cache-disabled medians; Supplementary Table S8 reports both settings.

Peak memory was the highest macOS process memory footprint (`phys_footprint`) during a run, sampled every 50 ms. Peak MLX active memory, the total size of live MLX buffers, is reported in Supplementary Table S8.

### Hardware and software

Package runtime and memory measurements and MLX output comparisons used a 2021 MacBook Pro with an M1 Max (10-core CPU, 32-core GPU) and 64 GB of unified memory. Network benchmarks used this computer and a MacBook Pro with an M5 Max (40-core GPU) and 128 GB of unified memory. The original implementations ran on the M1 Max CPU, the M5 Max CPU and an Intel Core i9-14900K desktop with an NVIDIA RTX 4090 GPU; Supplementary Table S3 gives the device for each single-case comparison. Both Macs ran Python 3.13 and MLX 0.32. The original implementations used PyTorch 2.13 or 2.14, MONAI 1.4 or 1.6, nnU-Net v2 2.8 and TensorFlow 2.20. Core ML exports used coremltools 9.0, and Core AI exports used the Core AI 1.0 beta.

### Runs of the original implementations

Comparisons used corresponding released checkpoints, prepared inputs, prompts and inference settings. Single-case runs of the original implementations used the M1 Max CPU, except TotalSegmentator and SAM 3, which used the Intel CPU, and LesionDiffusion, which used the RTX 4090 (Supplementary Table S3). Comparisons across multiple cases used the Intel CPU and RTX 4090 (Supplementary Table S7). LesionDiffusion was compared on the RTX 4090 only, because 200-step sampling was impractically slow on a CPU. NV-Reason-CT comparisons began after image preprocessing: the original network received the MLX-prepared CT, and its processor’s prompt tokens were checked against the MLX tokens. Supplementary Note 2 lists hardware and software adaptations.

Reference runs used FP32 by default, with automatic mixed precision and CUDA TF32 disabled. Model-specific exceptions retained the original arithmetic: FP16 sliding-window accumulation for nnU-Net, BF16 memory storage for MedSAM2, and FP32 LMNet with FP16 LINet and VQ for LesionDiffusion.

### Output agreement criteria

For segmentation masks, agreement required a Dice score of at least 0.99 for every label present in either output, including background. We also counted differing voxels. FleXray lung probability maps were thresholded at 0.5 for mask comparison. For both the single model and the five-model ensemble, all 61 probability channels, including background, were also compared on the five radiographs in Supplementary Table S5 across all three device pairs. Probability comparisons used whole-array and per-channel cosine similarity, mean absolute error and maximum absolute error, with the image region assessed separately from the padding.

Detections with scores of at least 0.02 were matched one to one using the Hungarian algorithm on box intersection over union (IoU). Agreement required equal detection counts and, for every matched pair, the same label, IoU of at least 0.99 and a score difference no greater than 0.001.

For SAM 3, each axial CT slice was selected by maximum annotated liver area and windowed to −150 to 250 HU. The text prompt was “liver” and the confidence threshold was 0.5. Predicted instances were matched by maximum mask IoU. Agreement required equal instance counts, per-instance mask IoU of at least 0.95, mean mask IoU of at least 0.99, a box-coordinate difference no greater than 2 pixels and a score difference no greater than 0.025. Agreement with the AMOS22 liver annotation was assessed separately; matching empty proposal sets counted as agreement between implementations.

Feature arrays and continuous-valued image outputs required whole-array cosine similarity of at least 0.9999; maximum absolute differences were also reported. Greedy text generation required identical tokens.

### Generative and additional device comparisons

Table 1 uses the published LesionDiffusion inference settings, termed source mode. Supplementary Figure S12 shows seven organ examples in conditioned mode, which enables phenotype-text conditioning with the released weights. Both modes and their case selection are described in Supplementary Note 3.

Generative comparisons used identical initial and per-step Gaussian noise, exported from MLX and supplied to the original code, so that framework-specific random-number generators did not affect the comparison. NV-Generate used five T1 image-mode seeds (0–4) and 30 rectified-flow steps. LesionDiffusion source mode used one 160 × 160 × 128 AMOS22 liver crop, seeds 23– 27 and 200 DDIM steps. Conditioned mode used the seven organ crops shown in Supplementary Figure S12, 200 steps and seed 23. Regional image errors were measured inside and outside the union of the lesion masks (Supplementary Table S6).

For the M5 Max renal comparison, the five CPTAC-CCRCC cases underwent the same registration, denoising and kidney cropping as the other renal comparisons. MLX ran on the M5 Max GPU and the original MONAI implementation on its CPU, both in FP32. Masks were compared voxel by voxel.

### Higher-precision arithmetic

Separate higher-precision profiles for renal SegResNet and SynthSeg 2.0 were evaluated on the same five cases as their standard profiles, across all three device pairs. Inputs, checkpoint values and agreement criteria were unchanged.

The renal profile (`compensated-fp48`) represented intermediate values in MLX hidden layers as pairs of FP32 numbers, combined overlapping predictions in FP64 on the CPU and exported FP32 logits; its MONAI reference [6] used FP64. The SynthSeg profile (`compensated-fp32`) used compensated arithmetic within the segmentation and parcellation operators, carrying the rounding error of each operation forward as a correction. Its inputs, weights and operator outputs remained FP32. Its TensorFlow reference used FP64 internal arithmetic with FP32 operator outputs. Both SynthSeg implementations retained FP32 quality-control networks.

Smoothing preceded left–right averaging, matching the original operation order. Supplementary Note 5 reports results by case.

### Network benchmarks with Core ML and Core AI

We compared 17 FP32 networks across MLX, Core ML and Core AI on the M1 Max and M5 Max. Networks were included when both their Core ML and Core AI exports passed the export accuracy check: the networks of 16 Table 1 models and the standalone SynthSeg 1.0 network. The other five Table 1 models were excluded: SynthSeg 2.0 and BioMedParse v2 lack complete exports, NV-Generate failed the export accuracy check, and LesionDiffusion and NV-Reason-CT have only partial exports.

Each runtime received the same saved synthetic image tensor with batch size one; input sizes are listed in Supplementary Table S9. SegResNet BraTS used a four-channel tensor spanning the full 240 × 240 × 160 volume rather than a sliding-window patch. Lung nodule timing covered six classification and box-regression head maps, excluding anchor decoding and nonmaximum suppression. SynthSeg 1.0 was evaluated as a standalone network, separately from the full SynthSeg 2.0 pipeline. SwinUNETR used the MONAI 1.6 graph in all runtimes, and VISTA-3D computed prompt embeddings on every call. SAM 3 used a 1008 × 1008 image, a 32-token text prompt and an attention mask; its export includes the original image model’s empty-geometry token and was checked against the Transformers geometry path with a fully masked box.

MLX used its GPU backend. The nnU-Net liver and TotalSegmentator networks used the `torch-order-fp32` profile, which follows PyTorch’s CPU accumulation order, and the nnU-Net pancreas and BraTS networks used native arithmetic. Core ML used CPU_AND_GPU, and Core AI used its GPU profile.

Each network and runtime combination was tested in five runs, each in a separate Python process. Each run performed one initial call, three warm-up calls and ten timed calls. The value for each run was the median of the ten timed calls, and the reported network time was the median of the five run medians. First-call time was the median of the five initial calls; model loading was timed separately and excluded. Timing began with the NumPy input and ended when the NumPy output was available, including data transfer. Outputs were checked against the original network outside the timed interval: they had to match its shape and data type, contain only finite values and differ from it by no more than an absolute tolerance of 0.001 to 0.1, depending on the network.

Before each run, the benchmark either waited for three consecutive samples with idle CPU and GPU and no thermal or performance warnings, or recorded the CPU, GPU and thermal state at the start of the run. The GPU idle threshold was 10% on the M1 Max and 5% on the M5 Max.

On the M5 Max, MLX TF32 arithmetic was disabled (`MLX_ENABLE_TF32=0`). For SegResNet BraTS, nnU-Net pancreas and the lung nodule detector, the Core ML and Core AI exports computed group and instance normalization statistics in stages of at most 1,024 values; these exports met the same tolerances against the original networks.

### Data

We used publicly available data to assess agreement between implementations and to illustrate model outputs. The supplement lists the data sources, case identifiers, model settings and licenses.

CPTAC-CCRCC: Data used in this publication were generated by the National Cancer Institute Clinical Proteomic Tumor Analysis Consortium (CPTAC). No patient volumes are redistributed.

## Data Availability

Model packages, benchmark code, software environments, evaluation results and instructions for obtaining the example cases will be available on GitHub. Supplementary Section S1 lists the evaluated packages, checkpoints, settings and input cases. Checkpoint terms and conversion instructions accompany the model packages. Evaluation datasets are available from their original providers.

https://github.com/MedMLX

https://huggingface.co/MedMLX

## Author contributions

Joseph Sandoval and Alfredo Sandoval Jr. contributed equally to model implementation, experiments and benchmarking, comparisons with the original code, data analysis and interpretation, figure preparation, software release and manuscript preparation. Alfredo Sandoval Jr. directed the scientific work, including the research questions, model selection and evaluation design. Arsalan Saleem supervised the project.

## Code and data availability

Model packages, benchmark code, software environments, evaluation results and instructions for obtaining the example cases will be available on GitHub. Supplementary Note 1 lists the evaluated packages, checkpoints, settings and input cases. Checkpoint terms and conversion instructions accompany the model packages. Evaluation datasets are available from their original providers.

## Ethics

This study used only publicly available, de-identified imaging data, and ethics committee approval was not required.

## Use of AI tools

Claude (Anthropic) was used to improve the readability of the manuscript. The authors reviewed and edited the text and take full responsibility for its content.

