## Supplemental Materials for "MedMLX: On-Device Medical Imaging AI for Apple Silicon"

### Supplementary Note 1. Models, checkpoints and datasets

The MCP server, started with `medmlx serve`, runs locally over standard input and output and does not require a network service. Each model call creates a job that the client can poll or cancel; disconnecting the client cancels its jobs.

**Table S1.** Evaluated models, checkpoints, settings and input cases. The identifier below each model name is the runner passed to `medmlx run`. A dash denotes the released default settings.

| Model | Package | Checkpoint | Settings | Input case |
|---|---|---|---|---|
| nnU-Net v2, liver<br>`nnunet` | nnunet-mlx | Zenodo 18807203 (liver and liver tumor), fold 0 | `3d_fullres` | AMOS22 amos_0008 [33] |
| nnU-Net v2, pancreas<br>`nnunet` | nnunet-mlx | Zenodo 17212158 (PancCTMultiTalentV2), `fold_all` | `3d_fullres_bs4` | MSD pancreas_052 [34] |
| nnU-Net v2, BraTS<br>`nnunet` | nnunet-mlx | Zenodo 11582627 (BraTS 2019), fold 0 | `3d_fullres` | MSD BRATS_001 (T1w, T1gd, T2w, FLAIR) [34], [35] |
| nnU-Net v2, autoPET<br>`nnunet` | nnunet-mlx | Zenodo 8362371 (autoPET II), fold 0 | `3d_fullres_resenc_bs80` | autoPET AP_16cdbc8689-0 (CT, SUV PET) [36], [37] |
| TotalSegmentator fast (3 mm)<br>`totalsegmentator` | totalsegmentator-mlx | TotalSegmentator v2, Dataset297, fold 0 | Fast (3 mm), 117 classes | AMOS22 amos_0008 [33] |
| SegResNet, BraTS<br>`segresnet` | segresnet-mlx | MONAI brats_mri_segmentation | — | MSD BRATS_001 (T1gd, T1w, T2w, FLAIR) [34], [35] |
| SegResNet, renal CECT<br>`segresnet` | segresnet-mlx | MONAI renalStructures_CECT_segmentation | — | CPTAC-CCRCC C3L-01458 (arterial, venous, excretory) [38], [39] |
| UNesT, renal<br>`unest` | unest-mlx | MONAI renalStructures_UNEST_segmentation | — | AMOS22 amos_0041 [33] |
| SwinUNETR, BTCV<br>`swin-unetr` | swin-unetr-mlx | MONAI swin_unetr_btcv_segmentation | — | AMOS22 amos_0008 [33] |
| SynthSeg 2.0<br>`synthseg` | synthseg-mlx | SynthSeg 2.0 segmentation, parcellation and QC | Standard (not robust) | MSD BRATS_001 (T1w) [34], [35] |
| VISTA-3D / NV-Segment-CT<br>`nv-segment-ct` | mlx-segment-ct | NV-Segment-CT (VISTA-3D) | Automatic mode; original nearest-label restoration | AMOS22 amos_0008 [33] |
| MedSAM2, CT lesion<br>`medsam2` | medsam2-mlx | MedSAM2_CTLesion | Box prompt; bidirectional propagation | LUNA16 …808059 [2], [40] |
| SAM 3<br>`sam3` | sam3-mlx | SAM 3 image detector, SAM License | Liver text prompt; 1008 pixels; confidence threshold 0.5 | AMOS22 amos_0013 (selected axial slice) [33] |

| Model | Package | Checkpoint | Settings | Input case |
|---|---|---|---|---|
| BioMedParse v2<br>`biomedparse-v2` | biomedparse-mlx | biomedparse_v2.ckpt | Existence threshold 0.5; spleen and right-kidney text prompts[a] | AMOS22 amos_0018 [33] |
| Lung nodule detector<br>`lung-nodule` | lung-nodule-mlx | MONAI lung_nodule_ct_ detection 0.6.10 | Raw-input resampling | LUNA16 …808059 [2], [40] |
| NV-Generate-MR-Brain<br>`nv-generate` | nv-generate-mlx | NV-Generate-MR-Brain | Image mode, T1, 30 flow steps, seed 0 | None (synthetic image) |
| LesionDiffusion<br>`lesiondiffusion` | lesiondiffusion-mlx | LesionDiffusion LMNet, LINet and VQ (EMA) | Liver tumor inside the parenchyma, 30 × 30 × 20 mm box; 200 DDIM steps; seed 23 | AMOS22 amos_0008 (160 × 160 × 128 crop) [33] |
| NV-Reason-CT<br>`mlx-reason-ct` | MLX-Reason-CT | NV-Reason-CT 386b93e0 | Structured chest report; greedy decoding, up to 512 new tokens, thinking off | LUNA16 …821260 [2], [40] |
| RadImageNet<br>`radimagenet` | radimagenet-mlx | RadImageNet-ResNet50_notop.h5 | Feature extraction | AMOS22 amos_0008, axial slice 50 [33] |
| FleXray<br>`flexray` | flexray-mlx | FleXray flux0375 | Lung label, threshold 0.5, no test-time augmentation | Wikimedia Commons normal PA radiograph [41] |
| FlexibleUNet<br>`flexible-unet` | flexible-unet-mlx | MONAI endoscopic_ tool_segmentation | — | Kvasir-Instrument ckcu8ty6z00003b5yzfa ezbs5 [42] |

[a] BioMedParse v2 prompts were “CT imaging of the spleen within the abdomen” for label 1 and “Presence of the right kidney detected in abdominal CT images” for label 2.

**Table S2.** Dataset licenses.

| Dataset | License |
|---|---|
| AMOS22 | CC BY 4.0 |
| Medical Segmentation Decathlon / BraTS | CC BY-SA 4.0 |
| LUNA16 | CC BY 4.0 |
| LIDC-IDRI | CC BY 3.0 |
| AutoPET-RG-Lym | CC BY-NC 4.0 |
| autoPET-III-Lite | CC BY-NC 4.0 |
| CPTAC-CCRCC, version 14 | CC BY 4.0 |
| Kvasir-Instrument | Research and education; commercial use requires permission |
| Normal PA chest radiograph (Wikimedia Commons) | CC0 1.0 |
| KiTS23 | CC BY-NC-SA 4.0 |
| TCGA-ESCA | CC BY 3.0 |
| Montgomery County CXR | Dataset-specific terms |

### Supplementary Note 2. Comparisons with the original implementations

Table S3 reports output agreement for one example case or generation seed for each of the 21 models. Model-specific settings and changes to the original implementations are described below; settings already listed in Tables S1 and S3 are not repeated. Supplementary Note 5 extends the comparisons to multiple cases, devices and arithmetic profiles.

**Table S3.** Output agreement for one input case or generation seed per model. MLX outputs are compared with those of the original implementations on the devices shown. Segmentation rows report the minimum Dice score across labels and the number of differing voxels; FleXray and FlexibleUNet differences are counted in pixels.

| Model | Original code | Original device | Measure | Value | Output difference |
|---|---|---|---|---|---|
| nnU-Net v2, liver | nnU-Net v2 (PyTorch) | M1 Max CPU, FP32 (FP16 accumulation) | Minimum label Dice | 0.99994 | 40 voxels |
| nnU-Net v2, pancreas | nnU-Net v2 (PyTorch) | M1 Max CPU, FP32 (FP16 accumulation) | Minimum label Dice | 0.99997 | 6 voxels |
| nnU-Net v2, BraTS | nnU-Net v2 (PyTorch) | M1 Max CPU, FP32 (FP16 accumulation) | Minimum label Dice | 1 | Identical mask |
| nnU-Net v2, autoPET | nnU-Net v2 (PyTorch) | M1 Max CPU, FP32 (FP16 accumulation) | Minimum label Dice | 1 | Identical mask |
| TotalSegmentator fast (3 mm) | TotalSegmentator (PyTorch) | Intel CPU, FP32 (FP16 accumulation) | Minimum label Dice | 1 | Identical mask |
| SegResNet, BraTS | MONAI bundle (PyTorch) | M1 Max CPU, FP32 | Minimum label Dice | 1 | Identical mask |
| SegResNet, renal CECT | MONAI bundle (PyTorch) | M1 Max CPU, FP32 | Minimum label Dice | 1 | Identical mask |
| UNesT, renal[a] | MONAI bundle (PyTorch) | M1 Max CPU, FP32 | Minimum label Dice | 1 | Identical mask |
| SwinUNETR, BTCV | MONAI bundle (PyTorch) | M1 Max CPU, FP32 | Minimum label Dice | >0.99999 | 1 voxel |
| SynthSeg 2.0 | SynthSeg (TensorFlow) | M1 Max CPU, FP32 | Minimum label Dice | 1 | Identical mask |
| VISTA-3D / NV-Segment-CT | NV-Segment-CT, MONAI (PyTorch) | M1 Max CPU, FP32 | Minimum label Dice | 1 | Identical mask |
| MedSAM2, CT lesion | MedSAM2 (PyTorch) | M1 Max CPU, FP32 (BF16 memory) | Minimum label Dice | 1 | Identical mask |
| SAM 3 | SAM 3, Transformers (PyTorch) | Intel CPU, FP32 | Instance proposal agreement | Pass | Mask IoU 0.99226; maximum box error 0.18201 pixels; maximum score difference 0.0080196 |
| BioMedParse v2[d] | Microsoft BioMedParse (PyTorch) | M1 Max CPU, FP32 | Maximum logit difference | $1.17 \times 10^{-3}$ | Identical mask |

| Model | Original code | Original device | Measure | Value | Output difference |
|---|---|---|---|---|---|
| Lung nodule detector | MONAI bundle (PyTorch) | M1 Max CPU, FP32 | Matched boxes | 7/7 | Minimum IoU >0.99999; maximum score difference 1.73 × $10^{-6}$ |
| NV-Generate-MR-Brain | NV-Generate, MONAI (PyTorch) | M1 Max CPU, FP32 | Image cosine | >0.99999 | — |
| LesionDiffusion[b] | LesionDiffusion (PyTorch) | RTX 4090, mixed precision | Image cosine | 0.99997 | Identical lesion mask |
| NV-Reason-CT | NV-Reason-CT, Transformers (PyTorch) | M1 Max CPU, FP32 | Generated tokens | Identical | — |
| RadImageNet | Keras ResNet50 (TensorFlow) | M1 Max CPU, FP32 | Minimum output cosine | >0.99999 | — |
| FleXray[c] | FleXray (PyTorch) | M1 Max CPU, FP32 | Minimum label Dice | 1 | Identical lung mask; 61-channel probability cosine >0.99999 |
| FlexibleUNet | MONAI bundle (PyTorch) | M1 Max CPU, FP32 | Minimum label Dice | 1 | Identical mask |

[a] UNesT, renal: the table uses amos_0041 because both implementations labeled the femoral heads as kidney on AMOS22 amos_0008 (Fig. S10). [b] LesionDiffusion: the original implementation ran LMNet in FP32 and LINet and VQ in FP16, with TF32 disabled. [c] FleXray mask agreement refers to the lung channel (59), thresholded at 0.5. [d] BioMedParse v2: MLX reused the random attention samples drawn by the original run.

#### *Model-specific settings and implementation changes*

**nnU-Net v2.** Comparisons used version 2.8.1, with mirroring disabled and a tile step of 0.5.

**nnU-Net v2, autoPET.** CT was aligned to the PET grid.

**TotalSegmentator fast (3 mm).** The reference was TotalSegmentator 2.12.0 running on the Intel CPU.

**SegResNet, BraTS.** Outputs used a sigmoid threshold of 0.5 and the BraTS label precedence 4 > 1 > 2.

**SegResNet, renal CECT.** The three CT phases were converted from DICOM, registered to the venous phase, cropped around the kidneys and denoised with median and curvature anisotropic diffusion filters. Both implementations received these prepared images. The MLX implementation reproduced the original FP32 convolution accumulation order and used Welford group normalization.

**UNesT, renal.** Inference used $96^3$ windows, sw_batch_size 4 and an overlap of 0.5.

**SwinUNETR, BTCV.** To fit in memory, the original implementation used sw_batch_size 1 instead of 4; all other sliding-window settings were unchanged.

**SynthSeg 2.0.** The original implementation included compatibility changes for tf_keras and NumPy. Comparisons included every output label, including the cortical parcels.

**VISTA-3D / NV-Segment-CT.** Inference used segment_all with 117 labels, $128^3$ patches and an overlap of 0.25.

**MedSAM2, CT lesion.** The box prompt was [411, 270, 442, 301] on slice 68, with bidirectional propagation over slices 60–76. The original CUDA-only connected-components step was replaced with an equivalent 8-connected OpenCV operation.

**Lung nodule detector.** Test-time augmentation was disabled.

**NV-Generate-MR-Brain.** Guidance was set to 2, and the original implementation received the initial noise generated in MLX. The original decoder used MONAI FP32 normalization because its FP16 option requires CUDA.

**LesionDiffusion.** Source-mode inference used a liver crop resampled to a 1 mm RAS grid. The original implementation received the MLX initial and per-step noise. MLX half-precision convolution reproduced PyTorch rounding. The original VQ decoder, which is hard-coded to a second GPU, was moved to the single RTX 4090.

**NV-Reason-CT.** The prompt tokens were verified to match across implementations, and the original network received the CT preprocessed by MLX.

**RadImageNet.** Comparisons included pooled and spatial features. The input slice was windowed to −160 to 240 HU and scaled to 8 bits.

**FleXray.** Outputs from the original InferenceRunner were mapped to the source image grid using the same procedure as the MLX package.

**FlexibleUNet.** Background and foreground outputs were compared at full resolution.

### Supplementary Note 3. Cases and settings for Figure 1 and Figure S12

Figure 1 presents selected examples of the model outputs. The liver, brain tumor and PET lesion cases had the highest whole-volume Dice scores against dataset annotations among five, five and three candidates, respectively. Liver-case selection used the combined liver and liver-tumor mask; brain-tumor and PET-lesion selection used all foreground labels. The UNesT case was selected for renal contrast enhancement. The SAM 3 case was selected because it produced a liver proposal at the fixed confidence threshold, and the displayed axial slice had the largest annotated liver area. All examples were reviewed visually. Table S4 identifies the model and case for each panel and records differences from the Table S1 settings.

**Table S4.** Models and input cases shown in Figure 1. Unless indicated otherwise, panels use the model and settings listed in Table S1.

| Panels | Model | Case | Relation to Table S1 |
|---|---|---|---|
| a, b | TotalSegmentator v2, fast (3 mm) | autoPET fdg_0117d7f11f CT | Not in Table S1; complete pipeline described below |
| c | SwinUNETR, BTCV | AMOS22 amos_0008 | Same case; identical output |
| d | VISTA-3D / NV-Segment-CT | AMOS22 amos_0008 | Same case; identical output |
| e | nnU-Net v2, liver | AMOS22 amos_0029 | Different case |
| f | UNesT, renal | AMOS22 amos_0013 | Different case, compared on all three devices |
| g | TotalSegmentator v2, standard (1.5 mm) | LUNA16 …536987 | Not in Table S1; mask identical to official TotalSegmentator on Intel CPU and RTX 4090 |
| h | Lung nodule detector | LUNA16 …808059 | Same case; same detections and top score |
| i, j | MedSAM2, CT lesion | LUNA16 …808059 | Same case and box; masks identical on all three devices |
| k | nnU-Net v2, autoPET | autoPET fdg_01140d52d8 | Different case |
| l | FleXray ensemble | Wikimedia Commons normal PA radiograph | Same radiograph, all five released models; the ensemble agreed in 15 of 15 device-pair comparisons (Table S7) |

| Panels | Model | Case | Relation to Table S1 |
|---|---|---|---|
| m | FlexibleUNet | Kvasir-Instrument ckcu8ty6z00003b5yzfaezbs5 | Same case; crop of the output from Table S1 |
| n | NV-Generate-MR-Brain | Synthetic image, seed 0 | Same seed |
| o, p | SynthSeg 2.0, segmentation and parcellation | Synthetic T1 from panel n | Different input |
| q | nnU-Net v2, BraTS | MSD BRATS_004 | Different case |
| r, s, t | SAM 3 | AMOS22 amos_0013 (axial slice) | Same case; selected for a nonempty liver proposal at threshold 0.5 among five cases |

#### *TotalSegmentator*

Figure 1a, b shows TotalSegmentator v2 fast-mode (3 mm) outputs for the CT from autoPET case `fdg_0117d7f11f`. The totalsegmentator-mlx package implements the complete workflow without PyTorch: image reorientation and resampling, inference in slabs, combination of segmentation labels from multiple models, restoration of labels to the input image grid, and export. The networks run on the Apple GPU through nnunet-mlx. The `torch-order-fp32` profile preserves PyTorch's CPU accumulation order in selected operations. We compared this workflow with official TotalSegmentator 2.12.0 running in PyTorch on the Intel CPU and RTX 4090.

For fdg_0117d7f11f, the fast-mode MLX mask was identical to the Intel CPU output and differed from the RTX 4090 output by one voxel (minimum per-label Dice 0.99993). The two original outputs differed at the same voxel. For AMOS22 `amos_0008`, fast and coarse modes produced identical masks on all three devices. In standard mode, the volume was divided into three slabs and predictions from five models were combined. The resulting MLX mask differed from each reference mask by nine voxels (minimum per-label Dice 0.999550); the Intel CPU and RTX 4090 masks were identical. Figure 1g shows standard-mode lung-lobe segmentation on LUNA16 …536987, for which the MLX mask was identical to both original outputs.

#### *LesionDiffusion*

LesionDiffusion was evaluated in two modes. Source mode follows the published inference settings, with phenotype-text conditioning disabled in LINet. Conditioned mode enables the LINet text-conditioning pathway using the released weights. Conditioned-mode comparisons used the original PyTorch networks with the same text conditioning enabled.

For the source-mode liver example in Table 1, the requested phenotype was hypodense, but the generated lesion had a mean attenuation of 73.4 HU, compared with 66.8 HU in nearby liver. Figure S11 compares this output with the conditioned-mode output.

Figure S12 shows seven conditioned-mode runs using the released checkpoints, 200 steps and seed 23. Each input covers a 240 mm region around the target organ, resampled to 1 mm. TotalSegmentator supplied the segmentations of the target organ and surrounding structures.

The bladder and stomach examples used KiTS23 [43], [44] case_00014; the colon, kidney and liver examples used KiTS23 case_00000. The lung example used LUNA16 …536987, and the esophageal example used TCGA-ESCA TCGA-VR-A8EY [45]. Cases, lesion locations, box sizes and phenotype prompts were selected after several trial runs. None of the generated lesions overlaps an annotated kidney tumor or lung nodule. TCGA-ESCA provides no lesion annotations, so the esophageal location was reviewed visually.

### Supplementary Note 4. Example outputs and observed failures

Figures S1–S7 show outputs from the models evaluated in Table 1. Figures S8–S11 document failures. On CPTAC-CCRCC C3L-01458, renal SegResNet labeled the vertebral body as neoplasm, the posterior spinal elements as artery and vein, and only part of the right kidney as parenchyma (Fig. S8). nnU-Net v2 undersegmented the liver on AMOS22 `amos_0008` (Fig. S9). On the same case, both UNesT implementations labeled the femoral heads as kidney and left the kidneys unlabeled (Fig. S10). In LesionDiffusion source mode, the generated liver lesion was brighter than the surrounding liver despite a hypodense prompt (Fig. S11; Supplementary Note 3).

#### *Example outputs*

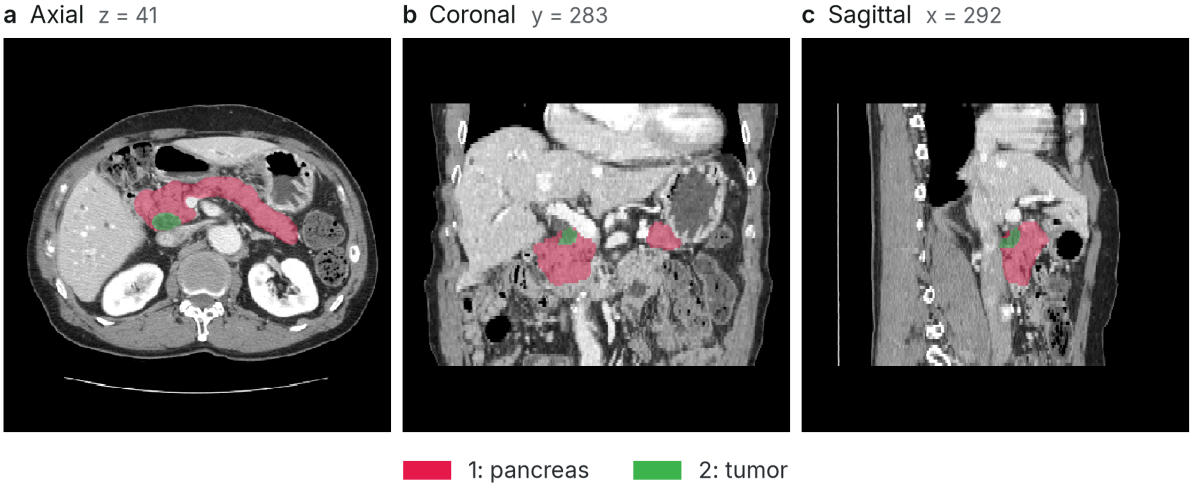


**Fig. S1.** Pancreatic segmentation with nnU-Net v2. Pancreas and pancreatic tumor labels on MSD pancreas_052.

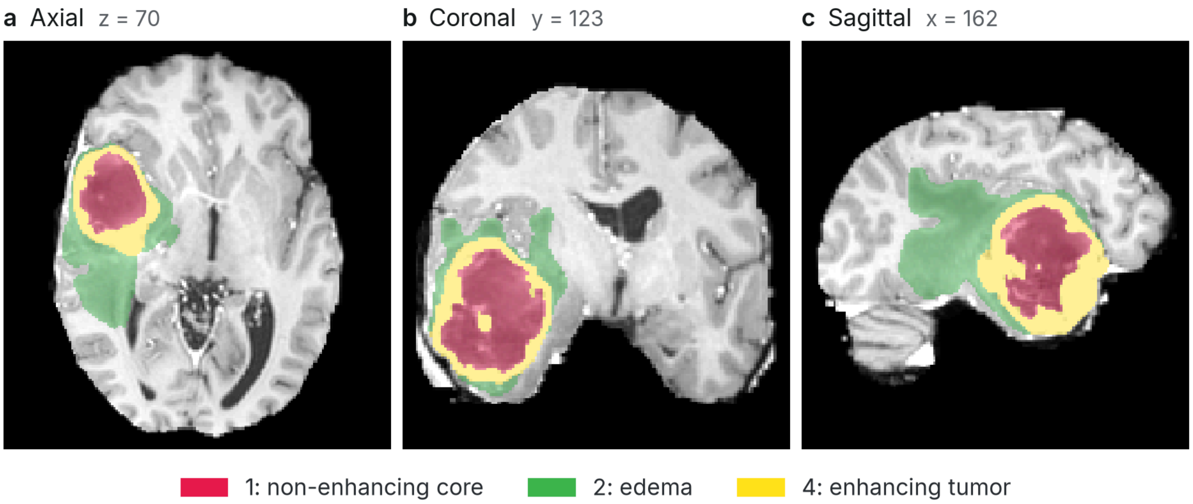


**Fig. S2.** Brain tumor segmentation with nnU-Net v2. Tumor subregion labels on MSD BRATS_001.

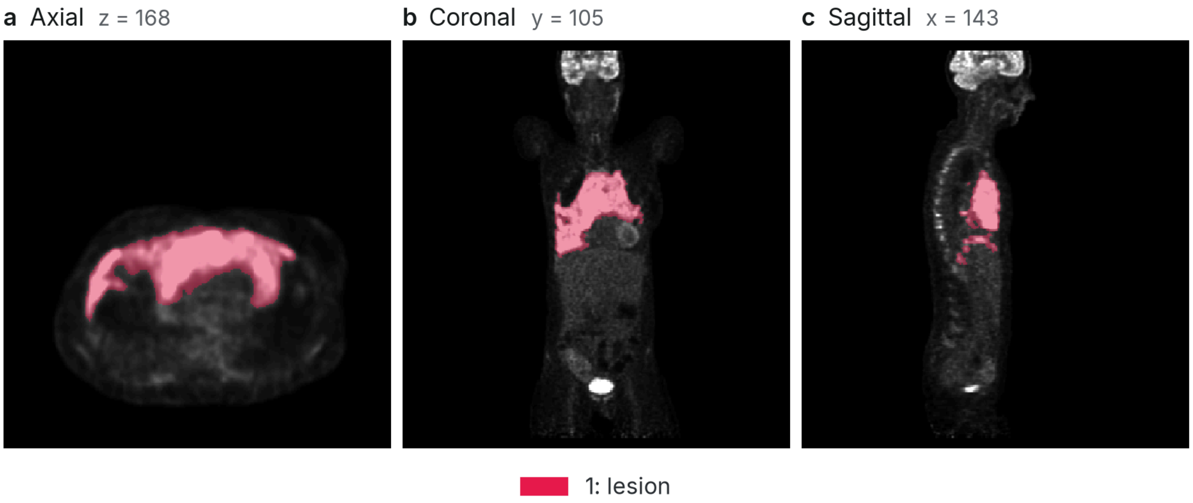


**Fig. S3.** FDG-avid lesion segmentation with nnU-Net v2. The lesion label is overlaid on the SUV PET image from autoPET AP_16cdbc8689-0.

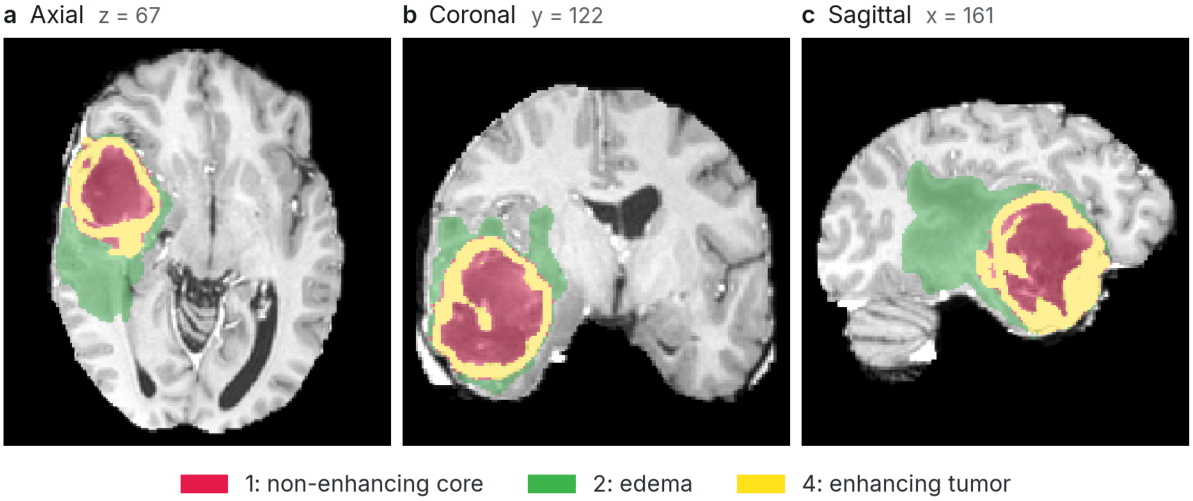


**Fig. S4.** Brain tumor segmentation with SegResNet. Tumor subregion labels on MSD BRATS_001.

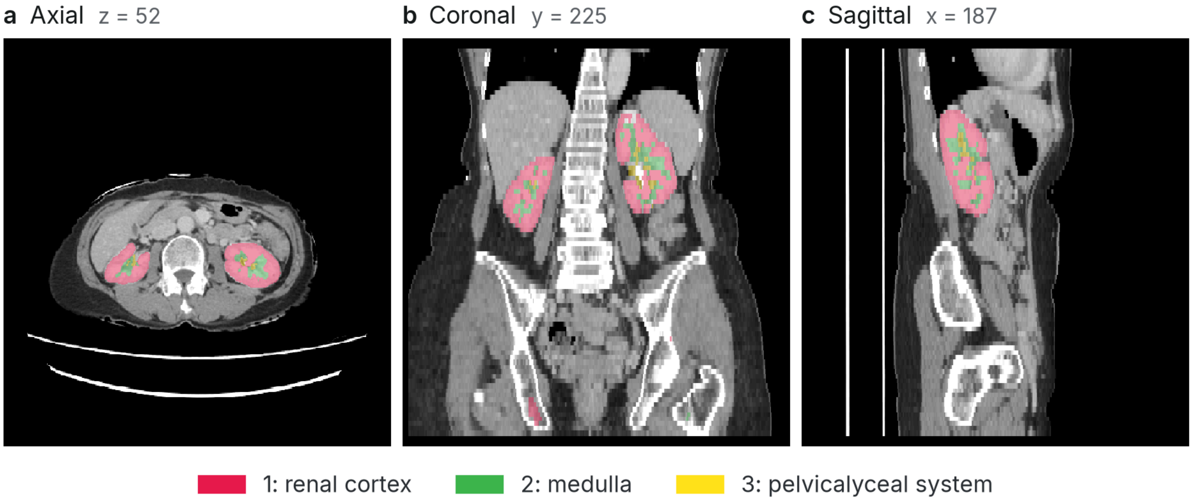


**Fig. S5.** Renal segmentation with UNesT. Renal cortex, medulla and pelvicalyceal system labels on AMOS22 amos_0041.

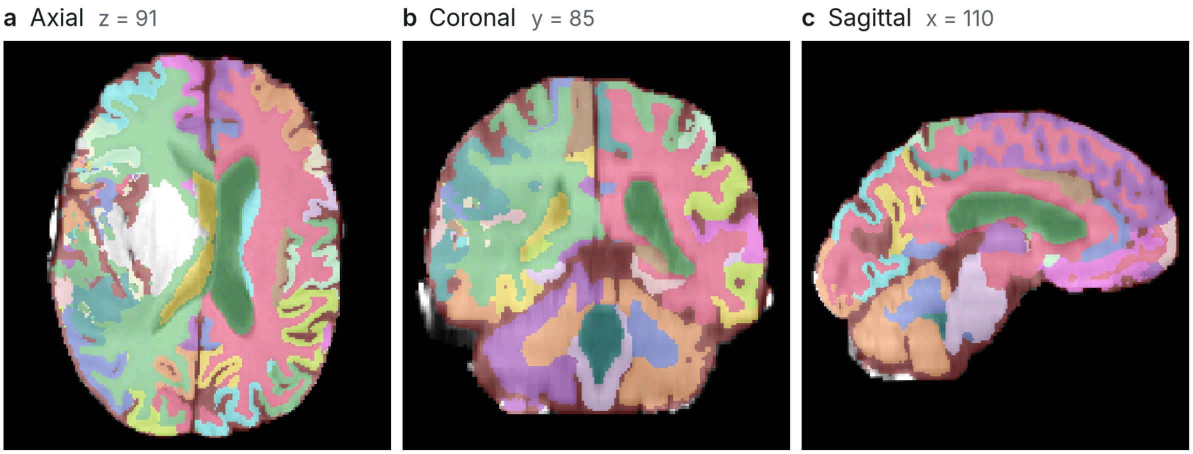


**Fig. S6.** Brain segmentation and parcellation with SynthSeg 2.0. The MSD BRATS_001 T1-weighted image is shown with 95 labels, including 68 cortical parcels. Colors distinguish neighboring labels. Quality-control scores ranged from 0.61 to 0.86.

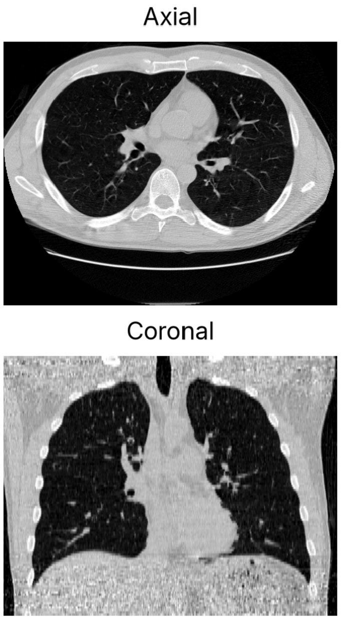


Generated report

TECHNIQUE: Non-contrast CT.

**FINDINGS:**
Medical devices: None present.

**CHEST:**
Lungs and airways: Trachea and main bronchi are patent. No active infiltration or mass lesion. A few tiny nonspecific nodules.
Pleura: No pleural effusion, pleural thickening, or pneumothorax.
Mediastinum and hila: No mediastinal lymphadenopathy or mass.
Heart and pericardium: Normal heart size. No pericardial effusion.
Great vessels: Normal caliber thoracic aorta and main pulmonary artery.
Esophagus: No pathological esophageal wall thickening.
Chest wall and breast: No chest wall or breast mass.
Bones: No lytic or destructive thoracic osseous lesion or acute fracture.

**IMPRESSION:**

1. A few tiny nonspecific pulmonary nodules.

**Fig. S7.** Chest CT report generation with NV-Reason-CT. Axial and coronal views of LUNA16 …821260 are shown alongside the 208-token structured report.

#### *Observed failures*

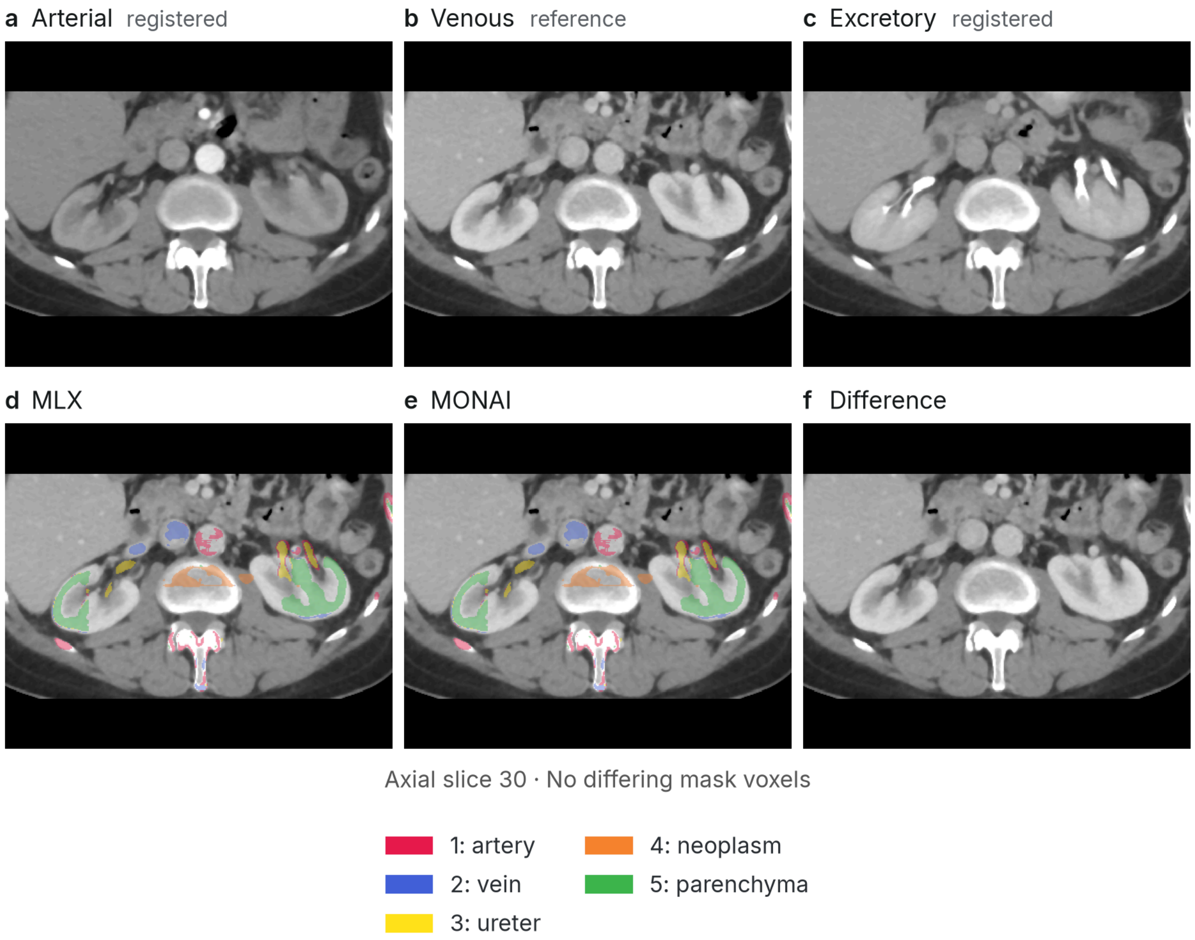


**Fig. S8.** Identical segmentation errors in MLX and MONAI renal SegResNet outputs. On CPTAC-CCRCC C3L-01458, both implementations label the vertebral body as neoplasm and the posterior spinal elements as artery and vein; only the lateral part of the right kidney is labeled as parenchyma. (a–c) CT phases after registration of the arterial and excretory phases to the venous phase. (d–f) MLX and MONAI masks overlaid on the venous phase and their difference.

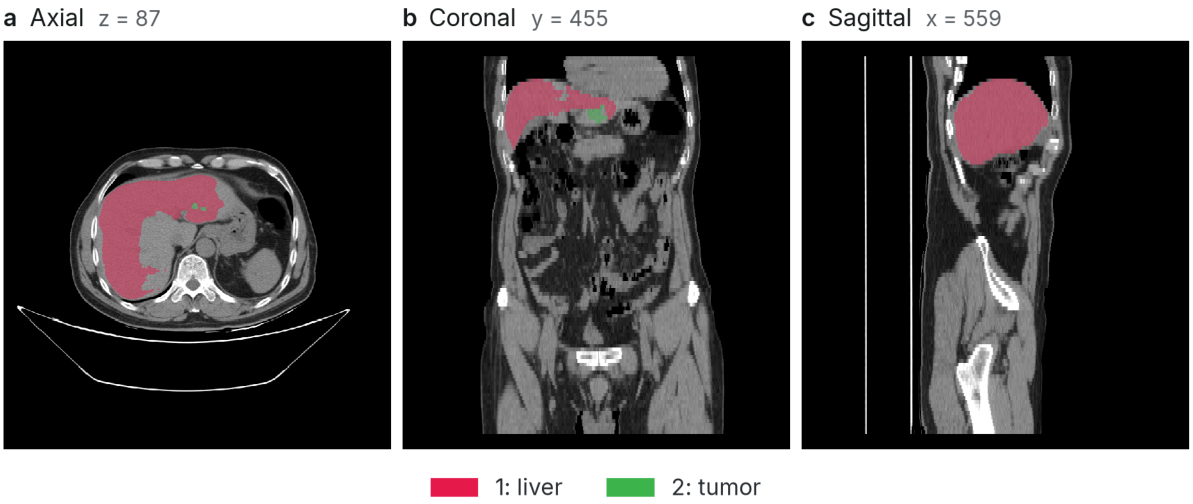


**Fig. S9.** Liver undersegmentation by nnU-Net v2. On AMOS22 amos_0008, the predicted liver mask has a Dice score of 0.76 against the dataset annotation.

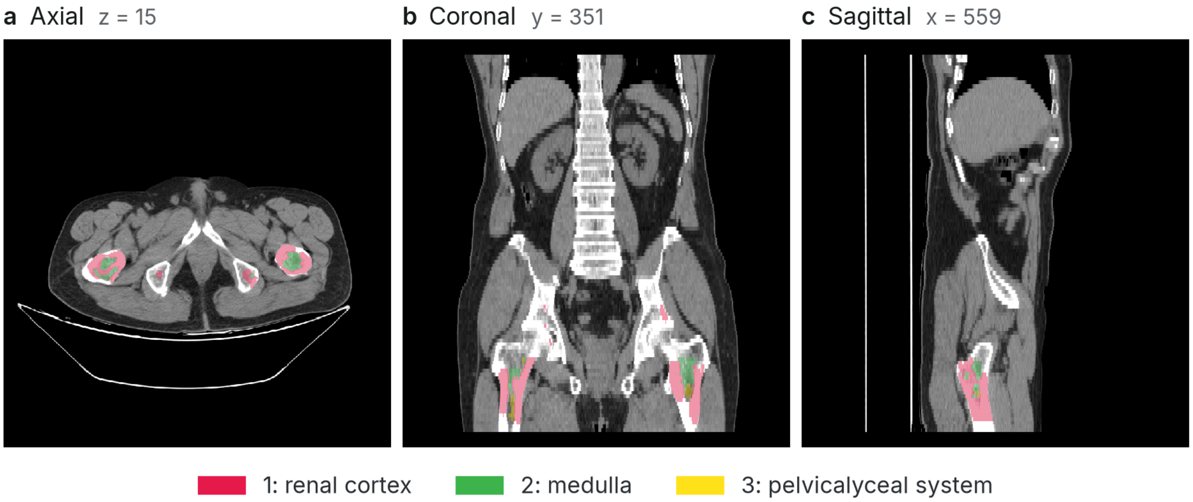


**Fig. S10.** Incorrect renal labels in both UNesT implementations. On AMOS22 amos_0008, the original and MLX implementations label the femoral heads as kidney and leave both kidneys unlabeled.

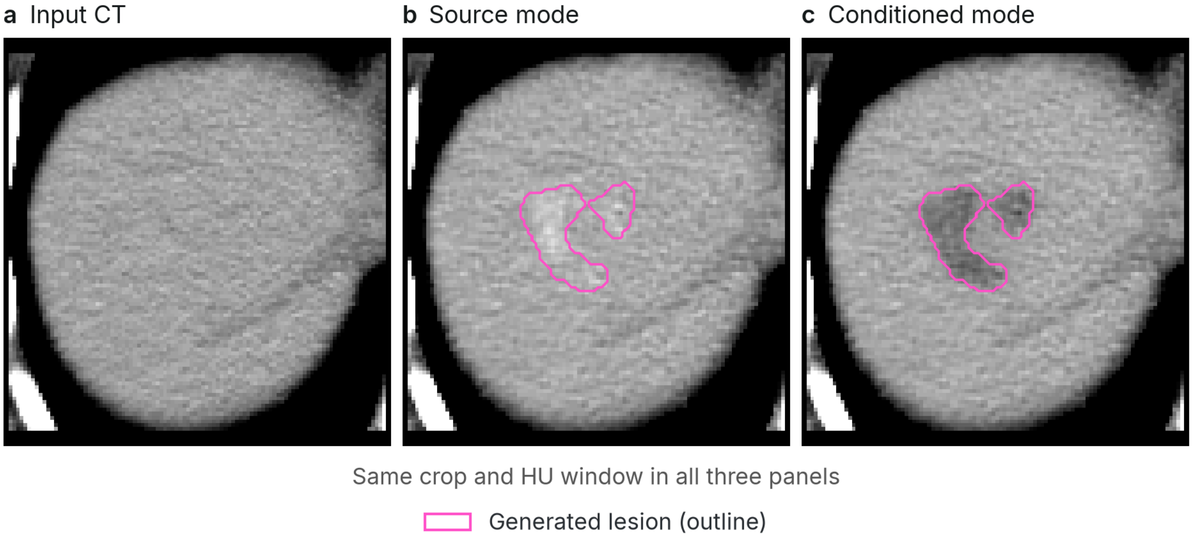


**Fig. S11.** Effect of phenotype conditioning on a LesionDiffusion example. For AMOS22 amos_0008, a hypodense liver lesion was requested. The source-mode output is brighter than the surrounding liver, whereas the conditioned-mode output is darker.

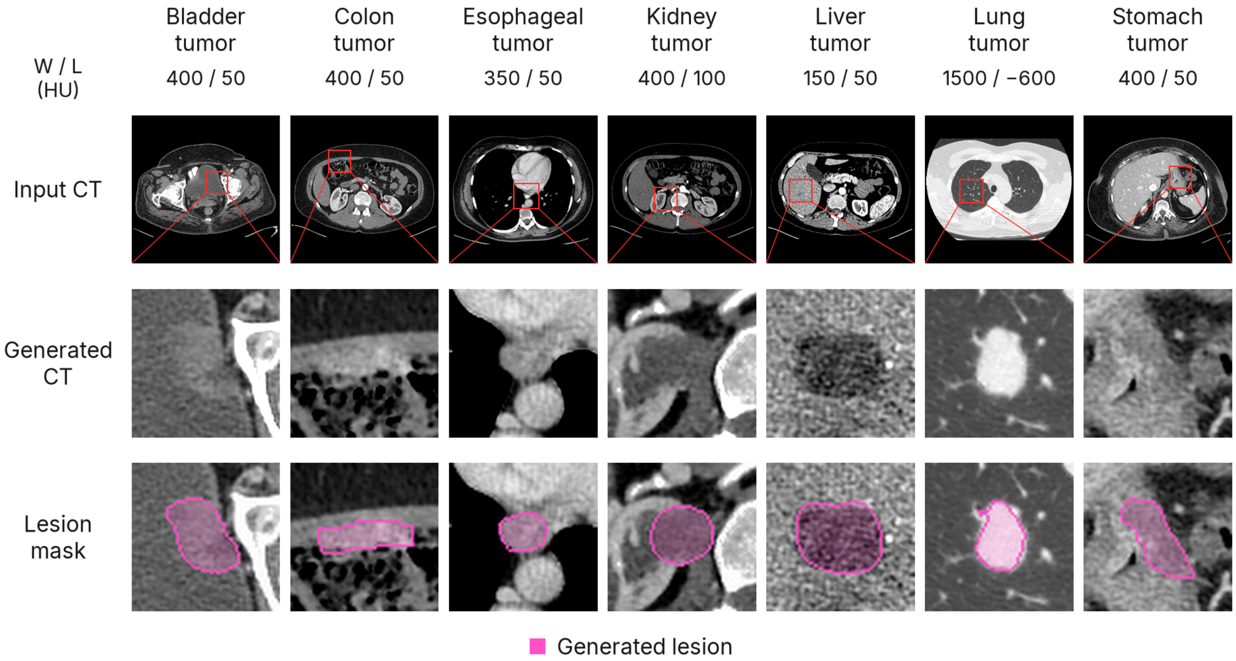


**Fig. S12.** Conditioned LesionDiffusion synthesis in seven organs. Top row: input CT images, with red boxes marking the enlarged regions. Middle row: the central axial slice from each generated volume. Bottom row: the same slice with the generated lesion mask in magenta. Window width and level (W/L, HU) are indicated above each column. TotalSegmentator [17] supplied the organ masks used for synthesis. Inputs comprise five KiTS23 examples [43], [44], LUNA16 …536987 [2] for the lung and TCGA-ESCA TCGA-VR-A8EY [45] for the esophagus. Lesion sites and phenotypes were selected after several trial runs. Table S6 reports comparisons with the original implementation using matched noise.

### Supplementary Note 5. Comparisons across cases, devices and arithmetic profiles

The standard-arithmetic evaluation comprised 324 model runs: 112 on the M1 Max GPU, 100 on the Intel CPU and 112 on the RTX 4090. Inputs came from 11 public imaging datasets. Each model or profile used five cases or generation seeds, except conditioned LesionDiffusion, which used one organ crop for each of seven tumor sites (Table S5). These comparisons covered 20 of the 21 models, all except BioMedParse v2, plus the FleXray ensemble and conditioned LesionDiffusion. The run counts exclude the higher-precision profiles and the repeat MLX runs of the Figure S12 examples, which are reported below. Some images were reused across generation seeds, organ crops and arithmetic profiles.

**Table S5.** Input cases and generation seeds for comparisons across devices. Models and profiles are grouped by shared inputs. LUNA16 identifiers are abbreviated to their final six digits, and autoPET entries use the study identifier. Counts include generation seeds and organ crops and therefore do not represent unique patients. Each renal input comprises three registered CT phases. The normal radiograph is the image used in Table S1.

| Models / profiles | Dataset / input | Cases or seeds |
|---|---|---|
| nnU-Net v2, BraTS<br>SegResNet, BraTS<br>SynthSeg 2.0, standard | MSD Task01 BrainTumour, training release | n = 5: `BRATS_001`, `BRATS_002`, `BRATS_003`, `BRATS_004`, `BRATS_005` |
| SwinUNETR, BTCV<br>VISTA-3D / NV-Segment-CT<br>nnU-Net v2, liver<br>UNesT, renal<br>RadImageNet<br>SAM 3 | AMOS22, validation release | n = 5: `amos_0008`, `amos_0013`, `amos_0018`, `amos_0022`, `amos_0029` |
| Lung nodule detector<br>NV-Reason-CT | LUNA16 subset0 | n = 5: `…821260`, `…741492`, `…808059`, `…536987`, `…656720` |
| NV-Generate-MR-Brain | T1 MRI synthesis; no patient image input | n = 5: `T1 seed 0`, `T1 seed 1`, `T1 seed 2`, `T1 seed 3`, `T1 seed 4` |
| FleXray<br>FleXray, five-model ensemble | Montgomery County CXR [46] and Wikimedia normal PA radiograph | n = 5: `MCUCXR_0001_0`, `MCUCXR_0002_0`, `MCUCXR_0104_1`, `MCUCXR_0108_1`, `Haggstrom-normal-PA-CC0` |
| FlexibleUNet | Kvasir-Instrument, test split | n = 5: `ckcu8ty6z00003b5yzfaezbs5`, `ckcu99qx200043b5yd2c5g84t`, `ckcu9j9vo00073b5y3m92mu6n`, `ckcu9jucf00083b5ytpqoue72`, `ckcu9ksoq00093b5ylw5wwfr2` |
| MedSAM2, CT lesion | LUNA16 subset0 | n = 5: `…741492`, `…808059`, `…536987`, `…874565`, `…755354` |
| nnU-Net v2, autoPET | autoPET FDG | n = 5: `fdg_0011f3deaf`, `fdg_01140d52d8`, `fdg_0117d7f11f`, `fdg_01682f60c3`, `fdg_0168f65af8` |
| nnU-Net v2, pancreas | MSD Task07 Pancreas, training release | n = 5: `pancreas_001`, `pancreas_004`, `pancreas_005`, `pancreas_006`, `pancreas_052` |
| LesionDiffusion | AMOS22 amos_0008 liver crop | n = 5: `amos_0008, seed 23`, `amos_0008, seed 24`, `amos_0008, seed 25`, `amos_0008, seed 26`, `amos_0008, seed 27` |

| Models / profiles | Dataset / input | Cases or seeds |
|---|---|---|
| TotalSegmentator 3 mm network | CPTAC-CCRCC and autoPET FDG | n = 5: C3N-00380, C3N-00435, fdg_0117d7f11f, fdg_01682f60c3, fdg_0168f65af8 |
| SegResNet, renal CECT, standard | CPTAC-CCRCC, registered three-phase CT crops | n = 5: C3L-01458, C3N-00380, C3N-00435, C3N-01524, C3N-01536 |
| LesionDiffusion, conditioned | KiTS23, LUNA16 and TCGA-ESCA | n = 7: liver: case_00000, bladder: case_00014, colon: case_00000, esophagus: TCGA-VR-A8EY, kidney: case_00000, lung: …536987, stomach: case_00014 |

#### *Scope and output agreement*

Where the complete pipeline was evaluated, comparisons included the model-specific processing through export; externally prepared inputs were shared between implementations as described in Supplementary Note 2. NV-Reason-CT comparisons began with the prepared CT and prompt tokens, and the five TotalSegmentator comparisons evaluated the network alone. Supplementary Note 3 reports separate comparisons of the complete TotalSegmentator pipeline. For SAM 3, two of the five selected slices produced no liver proposal on any device at the fixed confidence threshold of 0.5.

Of 312 standard-arithmetic comparisons, 310 met the agreement criteria: 211 of 212 comparisons between MLX and an original implementation, and 99 of 100 comparisons between the original implementations on the Intel CPU and RTX 4090. Both comparisons outside tolerance involved the Intel CPU output for SynthSeg 2.0 on BRATS_004 (Table S7); each had 55 differing voxels and a minimum per-label Dice score of 0.966667.

For FleXray, all 1,830 channel-level comparisons had cosine similarity above 0.9999, both over the full output grid and after exclusion of padding. The maximum absolute probability difference was $3.52 \times 10^{-5}$. These probability comparisons were evaluated separately from the lung-mask agreement counts in Table S7.

#### *Higher-precision profiles*

For the five renal cases in Table S5, the renal SegResNet `compensated-fp48` profile produced identical masks for all three device pairs.

The SynthSeg 2.0 `compensated-fp32` profile applies compensated arithmetic to convolution, normalization, activation, softmax and Gaussian smoothing. For the five brain MRI cases in Table S5, the masks were identical for all three device pairs.

#### *LesionDiffusion with matched noise*

Lesion masks were identical in all 12 comparisons with the original implementation on the RTX 4090 when the two implementations received the same noise (Table S6). Repeat MLX runs of each of the seven Figure S12 examples produced identical images and masks.

**Table S6.** LesionDiffusion output agreement with matched noise. MLX and the original implementation on the RTX 4090 used the same noise and 200 steps. Mask Dice compares the generated lesion masks. Image cosine similarity is calculated over the complete generated CT; lesion cosine similarity is calculated within the union of the two lesion masks. The final columns report mean and maximum absolute differences in HU inside and outside that union. Source-mode seeds 23–27 use the same AMOS22 liver crop. The seven conditioned organ examples are shown in Figure S12.

| Case | Mask Dice | Image cosine similarity | Lesion cosine similarity | Inside lesion: MAE / max (HU) | Outside lesion: MAE / max (HU) |
|---|---|---|---|---|---|
| amos_0008, seed 23 | 1.000000 | 0.99996977 | 0.99994968 | 0.507 / 4.883 | 0.775 / 261.719 |
| amos_0008, seed 24 | 1.000000 | 0.99996980 | 0.99996925 | 0.322 / 4.557 | 0.775 / 259.766 |
| amos_0008, seed 25 | 1.000000 | 0.99996977 | 0.99997679 | 0.389 / 5.208 | 0.776 / 259.766 |
| amos_0008, seed 26 | 1.000000 | 0.99996977 | 0.99993809 | 0.525 / 5.534 | 0.775 / 259.766 |
| amos_0008, seed 27 | 1.000000 | 0.99996979 | 0.99996637 | 0.334 / 4.883 | 0.776 / 259.766 |
| bladder: case_00014 | 1.000000 | 0.99999081 | 0.99993151 | 0.234 / 7.487 | 0.382 / 185.547 |
| colon: case_00000 | 1.000000 | 0.99999859 | 0.99993797 | 0.408 / 24.414 | 0.225 / 179.687 |
| esophagus: TCGA-VR-A8EY | 1.000000 | 0.99999371 | 0.99996749 | 0.385 / 7.650 | 0.625 / 210.938 |
| kidney: case_00000 | 1.000000 | 0.99999186 | 0.99994248 | 0.270 / 10.091 | 0.405 / 312.500 |
| liver: case_00000 | 1.000000 | 0.99999662 | 0.99991431 | 0.160 / 5.697 | 0.344 / 268.066 |
| lung: …536987 | 1.000000 | 0.99998990 | 0.99997665 | 0.579 / 38.086 | 0.701 / 428.223 |
| stomach: case_00014 | 1.000000 | 0.99999554 | 0.99995702 | 0.262 / 8.464 | 0.363 / 169.922 |

**Table S7.** Output agreement across devices and arithmetic profiles. Each cell gives the number of comparisons within tolerance divided by the total; a dash denotes an untested device pair. The final two rows report higher-precision profiles. For renal SegResNet, compensated-fp48 represents MLX hidden-layer values as pairs of FP32 numbers and is compared with FP64 MONAI references. For SynthSeg 2.0, compensated-fp32 uses compensated arithmetic with FP32 operator outputs and is compared with TensorFlow references using FP64 internal arithmetic (Methods). Cases include reused images, generation seeds and organ crops.

| Model / profile | Cases or seeds | M1 Max GPU–Intel CPU | M1 Max GPU–RTX 4090 | Intel CPU–RTX 4090 |
|---|---|---|---|---|
| nnU-Net v2, BraTS | 5 | 5/5 | 5/5 | 5/5 |
| SegResNet, BraTS | 5 | 5/5 | 5/5 | 5/5 |
| SwinUNETR, BTCV | 5 | 5/5 | 5/5 | 5/5 |
| VISTA-3D / NV-Segment-CT | 5 | 5/5 | 5/5 | 5/5 |
| nnU-Net v2, liver | 5 | 5/5 | 5/5 | 5/5 |
| UNesT, renal | 5 | 5/5 | 5/5 | 5/5 |
| SynthSeg 2.0, standard | 5 | 4/5 | 5/5 | 4/5 |
| Lung nodule detector | 5 | 5/5 | 5/5 | 5/5 |
| NV-Reason-CT | 5 | 5/5 | 5/5 | 5/5 |
| NV-Generate-MR-Brain | 5 | 5/5 | 5/5 | 5/5 |
| RadImageNet | 5 | 5/5 | 5/5 | 5/5 |
| FleXray | 5 | 5/5 | 5/5 | 5/5 |
| FleXray, five-model ensemble | 5 | 5/5 | 5/5 | 5/5 |

| Model / profile | Cases or seeds | M1 Max GPU–Intel CPU | M1 Max GPU–RTX 4090 | Intel CPU–RTX 4090 |
|---|---|---|---|---|
| FlexibleUNet | 5 | 5/5 | 5/5 | 5/5 |
| MedSAM2, CT lesion | 5 | 5/5 | 5/5 | 5/5 |
| nnU-Net v2, autoPET | 5 | 5/5 | 5/5 | 5/5 |
| nnU-Net v2, pancreas | 5 | 5/5 | 5/5 | 5/5 |
| LesionDiffusion | 5 | — | 5/5 | — |
| TotalSegmentator 3 mm network | 5 | 5/5 | 5/5 | 5/5 |
| SegResNet, renal CECT, standard | 5 | 5/5 | 5/5 | 5/5 |
| LesionDiffusion, conditioned | 7 | — | 7/7 | — |
| SAM 3 | 5 | 5/5 | 5/5 | 5/5 |
| SynthSeg 2.0, compensated-fp32 | 5 | 5/5 | 5/5 | 5/5 |
| SegResNet, renal CECT, compensated-fp48 | 5 | 5/5 | 5/5 | 5/5 |

### *Package measurements and network timings*

**Table S8.** Package runtime and memory for the workloads in main-text Table 1 on the M1 Max. With the default cache, runtime is the median (range) of five runs, and memory is the median of the five per-run peaks. With the cache disabled, values are medians of three runs. Peak memory is the process memory footprint; peak MLX active memory is the total size of live MLX buffers. Measurement settings are described in Methods.

| Model | Time (s) Default cache | Time (s) Cache disabled | Peak MLX active memory (GB) Default cache | Peak memory (GB) Default cache | Peak memory (GB) Cache disabled |
|---|---|---|---|---|---|
| nnU-Net v2, liver | 212.5 (204.0–218.0) | 263.2 | 5.4 | 17.8 | 6.1 |
| nnU-Net v2, pancreas | 33.9 (33.8–34.2) | 42.6 | 9.8 | 17.3 | 8.9 |
| nnU-Net v2, BraTS | 4.3 (4.3–4.4) | 4.9 | 3.1 | 5.5 | 3.2 |
| nnU-Net v2, autoPET | 13.9 (13.8–14.2) | 16.0 | 3.2 | 7.0 | 3.0 |
| TotalSegmentator fast (3 mm) | 3.0 (3.0–3.0) | 3.4 | 5.9 | 7.7 | 4.8 |
| SegResNet, BraTS | 1.3 (1.3–6.2) | 1.6 | 5.1 | 12.3 | 4.4 |
| SegResNet, renal CECT | 9.1 (9.0–9.7) | 10.8 | 4.0 | 9.5 | 4.5 |
| UNesT, renal | 104.1 (102.9–106.6) | 144.1 | 7.2 | 17.5 | 9.9 |
| SwinUNETR, BTCV | 88.4 (87.6–90.0) | 120.0 | 7.2 | 28.9 | 11.9 |
| SynthSeg 2.0 | 17.5 (17.1–17.9) | 17.4 | 11.6 | 14.2 | 12.2 |
| VISTA-3D / NV-Segment-CT | 44.9 (44.8–45.1) | 49.8 | 5.9 | 19.1 | 11.7 |
| MedSAM2, CT lesion | 3.0 (2.9–3.2) | 3.2 | 1.3 | 4.8 | 3.2 |
| SAM 3 | 1.7 (1.7–2.5) | 1.7 | 7.6 | 11.6 | 5.5 |
| BioMedParse v2 | 13.8 (13.4–14.2) | 14.3 | 2.9 | 4.9 | 2.9 |
| Lung nodule detector | 12.0 (11.6–12.1) | 11.8 | 5.7 | 6.8 | 6.7 |
| NV-Generate-MR-Brain | 251.2 (243.5–264.6) | 244.2 | 27.3 | 53.0 | 27.6 |

| Model | Time (s)<br>Default cache | Time (s)<br>Cache disabled | Peak MLX active memory (GB)<br>Default cache | Peak memory (GB)<br>Default cache | Peak memory (GB)<br>Cache disabled |
|---|---|---|---|---|---|
| LesionDiffusion | 1236.9 (1236.2–1577.4) | 1353.4 | 18.7 | 26.0 | 11.3 |
| NV-Reason-CT | 68.2 (67.4–73.2) | 84.8 | 22.7 | 53.5 | 23.3 |
| RadImageNet | 0.3 (0.2–0.3) | 0.2 | 0.1 | 0.3 | 0.3 |
| FleXray | 0.6 (0.6–1.1) | 1.1 | 1.9 | 2.6 | 1.1 |
| FlexibleUNet | 0.2 (0.2–0.3) | 0.2 | 0.7 | 1.2 | 0.4 |

**Table S9.** Controlled network timings on the M1 Max and M5 Max for Figure 4. Values are in milliseconds as the median (range) of five run medians; each run median summarizes ten calls after warm-up. Input dimensions exclude the batch and channel axes. SAM 3 also receives a 32-token prompt and an attention mask. Benchmark settings are described in Methods.

| Network | Input size | Computer | MLX | Core ML | Core AI |
|---|---|---|---|---|---|
| RadImageNet | 224 × 224 | M1 Max | 10.7<br>(10.7–17.4) | 4.0<br>(3.9–4.0) | 6.3<br>(6.2–6.4) |
| | | M5 Max | 6.0<br>(5.9–6.0) | 2.0<br>(2.0–2.1) | 2.9<br>(2.9–3.4) |
| MedSAM2, CT lesion | 512 × 512 | M1 Max | 21.3<br>(21.1–21.6) | 20.4<br>(20.3–26.4) | 29.7<br>(29.3–31.2) |
| | | M5 Max | 10.1<br>(10.0–10.1) | 8.8<br>(8.6–8.8) | 12.7<br>(12.6–13.1) |
| FlexibleUNet | 736 × 480 | M1 Max | 28.7<br>(28.2–28.9) | 13.3<br>(13.2–13.5) | 20.4<br>(20.3–20.9) |
| | | M5 Max | 14.0<br>(13.9–14.6) | 6.5<br>(6.1–8.2) | 11.0<br>(10.3–11.9) |
| SynthSeg 1.0 | $64^3$ | M1 Max | 43.9<br>(38.3–44.4) | 65.8<br>(61.4–66.9) | 126.2<br>(121.0–127.3) |
| | | M5 Max | 17.7<br>(17.6–17.8) | 28.6<br>(27.8–28.8) | 49.7<br>(49.5–50.0) |
| FleXray | 256 × 256 | M1 Max | 69.8<br>(68.8–71.1) | 26.1<br>(25.7–26.7) | 27.0<br>(26.9–28.8) |
| | | M5 Max | 32.7<br>(32.1–34.9) | 14.7<br>(14.7–14.9) | 16.0<br>(15.6–16.2) |
| TotalSegmentator fast (3 mm) | 112 × 112 × 128 | M1 Max | 135.3<br>(128.9–154.2) | 599.5<br>(543.4–614.3) | 941.7<br>(927.8–947.7) |
| | | M5 Max | 79.2<br>(78.9–79.4) | 244.3<br>(243.4–245.1) | 314.9<br>(314.2–315.0) |
| SegResNet, renal CECT | $96^3$ | M1 Max | 112.1<br>(111.7–113.0) | 294.6<br>(294.3–294.8) | 631.9<br>(630.2–633.8) |
| | | M5 Max | 79.6<br>(78.7–82.3) | 141.4<br>(132.3–167.2) | 439.5<br>(439.4–439.9) |
| UNesT, renal | $96^3$ | M1 Max | 175.7<br>(174.9–176.2) | 287.7<br>(287.6–288.0) | 470.7<br>(469.6–478.9) |
| | | M5 Max | 82.2<br>(82.0–82.2) | 129.8<br>(129.7–130.1) | 154.1<br>(152.7–154.2) |

| Network | Input size | Computer | MLX | Core ML | Core AI |
|---|---|---|---|---|---|
| SwinUNETR, BTCV | $96^3$ | M1 Max | 423.1 (421.0–424.0) | 486.0 (483.1–486.8) | 731.7 (730.2–735.2) |
| | | M5 Max | 155.6 (153.2–160.2) | 230.4 (226.0–337.7) | 285.9 (282.4–334.4) |
| nnU-Net v2, BraTS | 128 × 160 × 112 | M1 Max | 396.8 (386.5–400.8) | 658.3 (654.1–661.9) | 1,168.1 (1,163.0–1,219.4) |
| | | M5 Max | 178.7 (175.1–201.6) | 392.0 (384.3–405.3) | 457.6 (452.5–471.0) |
| nnU-Net v2, liver | $128^3$ | M1 Max | 393.7 (393.3–396.4) | 578.3 (578.2–578.4) | 975.6 (974.6–978.3) |
| | | M5 Max | 203.7 (202.5–205.5) | 260.1 (259.4–261.4) | 310.8 (310.0–312.8) |
| nnU-Net v2, autoPET | $128^3$ | M1 Max | 475.5 (472.9–485.0) | 848.8 (847.5–875.7) | 1,467.0 (1,461.7–1,491.4) |
| | | M5 Max | 207.1 (206.0–221.8) | 387.8 (382.9–419.3) | 452.8 (438.0–501.5) |
| SegResNet, BraTS | 240 × 240 × 160 | M1 Max | 430.2 (427.6–432.6) | 1,113.7 (1,112.8–1,117.5) | 2,275.2 (2,271.6–2,279.4) |
| | | M5 Max | 264.4 (260.2–266.7) | 528.0 (507.9–560.7) | 700.1 (694.8–731.8) |
| SAM 3 | 1008 × 1008 | M1 Max | 982.8 (977.9–997.5) | 960.0 (947.4–961.8) | 1,032.3 (1,019.2–1,033.8) |
| | | M5 Max | 529.0 (528.0–530.0) | 506.7 (500.6–663.0) | 531.3 (520.3–550.4) |
| VISTA-3D / NV-Segment-CT | $128^3$ | M1 Max | 1,035.5 (1,034.5–2,925.9) | 3,002.7 (2,986.8–3,026.3) | 5,064.4 (5,059.1–5,069.0) |
| | | M5 Max | 543.6 (525.4–2,092.9) | 1,409.7 (1,366.7–1,427.5) | 1,639.2 (1,628.7–1,690.0) |
| nnU-Net v2, pancreas | $192^3$ | M1 Max | 1,568.1 (1,554.7–1,579.8) | 2,852.0 (2,833.6–3,170.5) | 5,036.4 (5,033.0–5,333.8) |
| | | M5 Max | 706.0 (664.0–714.1) | 1,425.7 (1,257.0–1,447.2) | 1,416.2 (1,243.0–1,430.5) |
| Lung nodule detector | 256 × 256 × 128 | M1 Max | 1,788.9 (1,785.7–1,794.6) | 5,645.2 (5,640.1–5,662.3) | 10,742.0 (10,693.8–10,749.3) |
| | | M5 Max | 993.2 (912.8–1,001.3) | 2,642.6 (2,544.6–2,826.7) | 2,829.9 (2,601.2–3,003.7) |
